# An integrated framework for building trustworthy data-driven epidemiological models: Application to the COVID-19 outbreak in New York City

**DOI:** 10.1101/2021.02.22.21252255

**Authors:** Sheng Zhang, Joan Ponce, Zhen Zhang, Guang Lin, George Karniadakis

**Author notes:** Corresponding author: Guang Lin. Equal contribution (Sheng Zhang, Joan Ponce, Zhen Zhang).

## Abstract

Epidemiological models can provide the dynamic evolution of a pandemic but they are based on many assumptions and parameters that have to be adjusted over the time when the pandemic lasts. However, often the available data are not sufficient to identify the model parameters and hence infer the unobserved dynamics. Here, we develop a general framework for building a trustworthy data-driven epidemiological model, consisting of a workflow that integrates data acquisition and event timeline, model development, identifiability analysis, sensitivity analysis, model calibration, model robustness analysis, and forecasting with uncertainties in different scenarios. In particular, we apply this framework to propose a modified susceptible–exposed–infectious–recovered (SEIR) model, including new compartments and model vaccination in order to forecast the transmission dynamics of COVID-19 in New York City (NYC). We find that we can uniquely estimate the model parameters and accurately predict the daily new infection cases, hospitalizations, and deaths, in agreement with the available data from NYC’s government’s website. In addition, we employ the calibrated data-driven model to study the effects of vaccination and timing of reopening indoor dining in NYC.

## Introduction

This study aims to answer a fundamental question: given epidemiological data, how to develop an appropriate model and identify which parameters we can accurately infer that would, in turn, allow us to correctly predict the states of interest such as daily cases, hospitalizations, and deaths. The objective of this work is to provide a systematic way to model a pandemic accurately through carefully formulating a suitable model, uniquely identifying the model parameters, and predicting outbreaks under uncertainties based on the different epidemiological data available. To address the above fundamental question, we propose a general integrated framework to approach the problem systematically through identifiability analysis, sensitivity analysis, model robustness analysis, and forecasting under uncertainties.

Numerous modeling approaches have been used to gain insight into epidemic disease’s ever-evolving dynamics and the effects the interventions have had on containing the spread. Mathematical modeling is an efficient way to test and evaluate the effectiveness of hypothetical interventions that cannot be tested out due to practical or ethical limitations. Describing the disease’s development involves representing a highly complex process affected by social, environmental, and biological factors. Compartmental models have traditionally been used to depict systems that include individuals with different health statuses that change in time. In particular, mathematical modeling of COVID-19 using compartmental models described by ordinary differential equations (ODEs) such as susceptible–infectious–recovered (SIR) [1–3], modified SIR [1, 4–6], susceptible–exposed–infectious–recovered (SEIR) [7–9] and modified SEIR models [10, 11] has been used extensively in an attempt to capture the virus’ spread. These types of lumped mechanistic models, unlike data-driven models, can explore future outcomes of the pandemic and evaluate the effects of various interventions. Compartmental models are commonly applied in epidemiology for the reason that they are simple and easily tractable. However, these models are local and have limited capabilities to describe spatial dynamics. Partial differential equation (PDE) models, such as diffusion–reaction models, describe dynamics in time and space more naturally [12, 13]. ODE models are the most commonly used models due to the increased mathematical complexity and computational cost of PDE models. However, their accuracy is constrained by parameter uncertainties and gaps in information about the disease dynamics. Likewise, assumptions to maintain model simplicity may affect the estimated values. For a long-lasting pandemic, the model parameters change with time; hence the parameter identification problem becomes nontrivial given the fact that typically a limited amount of relevant data is available.

In an ODE-based epidemiological model, the system parameters usually contain critical information that often cannot be measured directly such as the transmission rate which needs to be inferred from data. A necessary condition for the well-posedness of a parameter estimation problem in ODE theory is the model’s structural identifiability if we assume noise-free data. The structural identifiability analysis can be performed without any experimental data; it addresses whether the parameter estimation problem is well-posed under ideal conditions. Should the postulated model not be structurally identifiable, the parameters obtained will be unreliable. However, a model can be structurally identifiable (a necessary condition) but may not be practically identifiable. Thus, the structural identifiability analysis may conclude that a model’s parameters are uniquely determined, yet when real-life, noisy data are used, the estimated parameter values could still be unreliable. To conduct the practical identifiability analysis, we compute the correlation matrices of model parameters in different settings using Fisher Information Matrices (FIMs) following lines of approach in [14, 15].

Non-identifiability is a problem frequently encountered in pandemics modeling since, typically, not every state variable is available. In recent literature, model identifiability issues have been studied due to the wide variation in model predictions in the context of the COVID-19 pandemic [2, 16–18]. Tuncer et al. analyzed the structural and practical identifiability of some of the most widely-used pandemic models, including SIR, SIR with treatment, and SEIR, assuming only one observable is available using simulated data [19]. Roda et al. extended these ideas by studying SIR and SEIR models’ practical identifiability using data from the COVID-19 outbreak in Wuhan, using only the counts of infected individuals as the available data [2]. They found that complex mechanistic models are more likely to have identifiability issues compared to simpler models. Massoni et al. provided a systematical structural identifiability and observability analysis of 255 available compartmental models for COVID-19 and found that approximately one-third of them have structurally non-identifiable parameters [16]. Therefore, an identifiability test should be conducted as a sanity check when a new ODE-based mechanistic model is proposed, to ensure trustworthy parameter estimation.

Furthermore, precise estimates of parameters allow us to determine an epidemiologically relevant value called the basic reproduction number, *ℛ*_0_. The basic reproduction number is defined as the average number of infections caused by one infected individual in an entirely susceptible population when disease control is absent, and it determines whether a disease will persist or not in a population.

Still, uncertainty about parameter values could be relatively high at the beginning of an outbreak even if identifiability is guaranteed. Therefore, determining the response of a model’s output to parameter variation helps identify sources of uncertainty. Sensitivity analysis studies how the uncertainty in the model’s output can be allocated to different inputs’ uncertainty sources [20]. It allows a better understanding of the model to analyze how the model parameters affect the output.

In the present work, we propose to integrate these steps of identifiability and sensitivity analysis together with policy change timelines, data availability, and uncertainties in forecasting. We apply the proposed general framework to introduce a modified SEIR model, which we extend to include vaccination, and forecast the transmission dynamics of COVID-19 in New York City (NYC) under vaccination and different safety measures relaxation scenarios. Daily cases, hospitalizations, and deaths in NYC are used to demonstrate the way to employ the proposed framework for simulating the ongoing COVID-19 pandemic in the city from early 2020 until February 2021.

COVID-19, which emerged in China in late 2019, has caused an outbreak affecting over 200 countries worldwide and was declared a pandemic on March 11, 2020 [21] by the World Health Organization. The strategies to control the spread of the virus in 2020 (before a vaccine was available) were mainly directed towards non-pharmaceutical interventions, such as isolation of infected individuals, social distancing, and face-mask use. COVID-19 was detected in a patient in NYC in early 2020 [22, 23]. The high population density and lack of control measures in the first three weeks produced an exponential increase in cases, which exceeded 20,000 before the statewide stay-at-home order was put in place on March 22, 2020 [24]. Between March 22, 2020, and June 8, 2020, a host of social distancing measures and non-essential businesses closings in NYC lowered the incidence from almost 2000 people infected a day in April to a couple of hundred cases by June 8, 2020 [25]. The city’s first phase of its four-phase reopening plan began on June 8, 2020, and the final stage started on July 20, 2020.

The proposed model considers presymptomatic, asymptomatic, hospitalized, isolated, and deceased individuals. Structural identifiability, practical identifiability, and sensitivity of the model are studied. Once the parameters that can be reliably estimated are identified, the parameter estimation portion of the study is broached. Then, the NYC outbreak data (daily infected, hospitalized, and deceased individuals) are employed to estimate the model parameters to understand how public policy such as isolation, public closings, and other social distancing measures impact the transmission dynamics. Moreover, confidence intervals are computed, and the model’s predictive capabilities are analyzed by considering the uncertainty of policies in constant flux. Next, we further investigate model robustness. The main modeling assumption is that the transmission rate is changing in time, following different policy measures, in a piecewise fashion. The vaccination deployment in NYC is incorporated into the model, and we evaluate the combined effect of vaccination and the gradual reopening process currently underway.

The novelty of this work is multi-fold:

- We develop a general framework and workflow for building a trustworthy data-driven epidemiological model;
- We propose a modified SEIR model with vaccination and validate it with the pandemic data in New York City (daily cases, hospitalizations, and deaths); every parameter in our model has physical meaning;
- We systematically study the structural identifiability, practical identifiability, and sensitivity to examine the relationship between the model dynamics, data, and parameters;
- We treat the transmission rate, hospitalization ratio, and death from hospital ratio as time-dependent model parameters based on the event-timeline, and calibrate the identifiable model parameters using simulated annealing and MCMC simulations. We also investigate model robustness by studying how the model behaves under random perturbations;
- We demonstrate the model’s predictive capabilities under uncertainties for different future scenarios;
- We specifically investigate the effects of indoor dining reopening and vaccination scenarios as a reference for policymakers.

### General Framework and Workflow

Holmdahl and Buckee, in [26], discussed different types of models for the COVID-19 epidemic as well as the distinct challenges in these approaches. The authors highlighted that “models are a way to formalize what we know about the viral transmission and explore possible futures of a system that involves nonlinear interactions, something that is almost impossible to do using intuition alone.” They further elaborate that “models will be useful for exploring possibilities rather than making strong predictions about longer-term disease dynamics.” Thus, a systematic way of designing an effective data-driven model is essential for assessing ongoing control strategies endowed with uncertainty quantification.

To systematically design an effective data-driven model, we propose a general framework for building a trustworthy data-driven epidemiological model, which constructs a workflow to integrate data acquisition and event timeline, model development, identifiability analysis, sensitivity analysis, model calibration, model robustness analysis, model prediction with uncertainties and investigation of reopening scenarios together. Fig 1 gives an overview of the framework, with details for each stage provided below.

**Fig 1.**
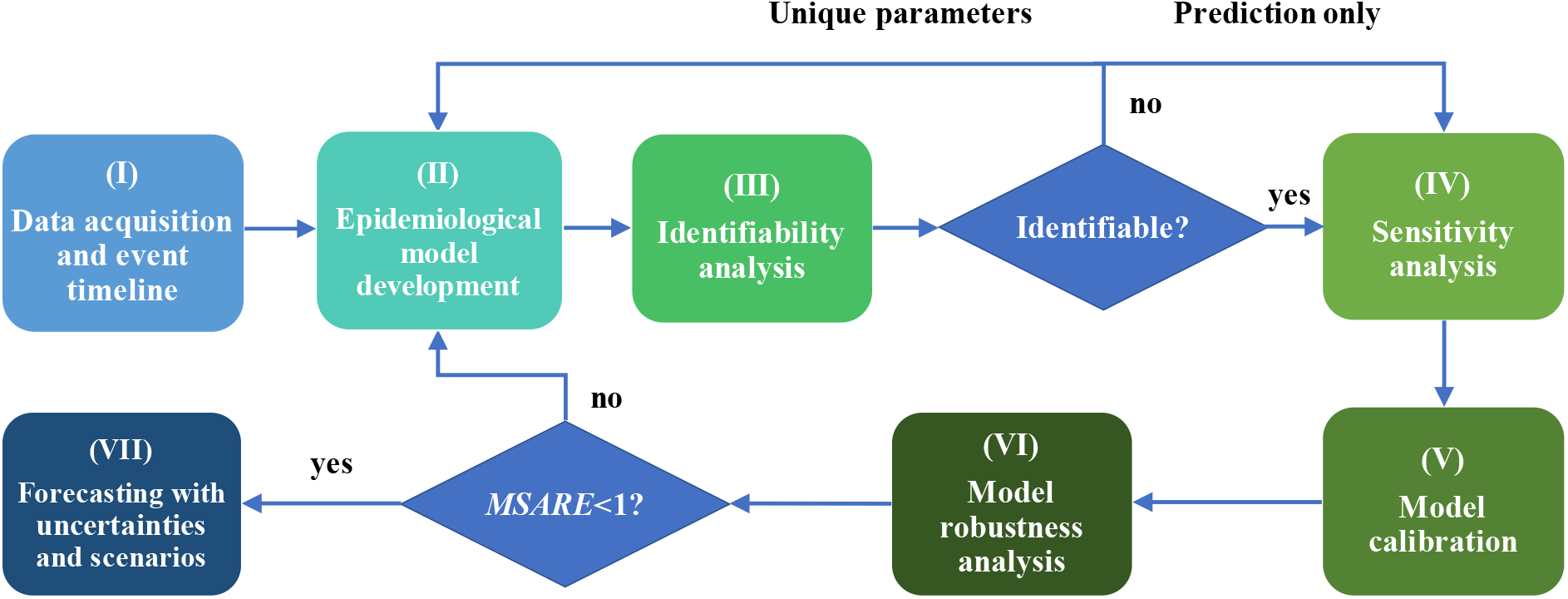
A general framework for building a trustworthy data-driven epidemiological model — an overview of the main contribution. In this work, we propose a general framework for building a trustworthy data-driven epidemiological model, which constructs a workflow to integrate data acquisition and event timeline, model development, identifiability analysis, sensitivity analysis, model calibration, model robustness analysis, and forecasting with uncertainties and scenarios. We first introduce a modified SEIR model that accommodates the pandemic data in New York City. Secondly, we study the structural identifiability, practical identifiability, and sensitivity to examine the relationship between the model’s data and parameters. We then calibrate the identifiable model parameters using simulated annealing and MCMC simulation. Model robustness is then checked to study how the model behaves under random perturbations. In addition, we demonstrate the model’s predictive capabilities with uncertainties. Finally, reopening scenarios are investigated as a reference for policymakers.

I. Acquire data and look for major interventions or events that could affect the transmission dynamics of the epidemic.
II. Develop an epidemiological model that accommodates the data and events in (I). The intervention events can be encoded through *time-dependent* parameters in the model.
III. Use both structural and practical identifiability analysis to determine the parameters to fit. Domain knowledge can also be incorporated in this step to help choose the parameters. If the model is not identifiable but one prefers unique parameters, one should fix some of the non-identifiable parameters in the model or propose other models. Otherwise, one can proceed to (IV).
IV. Conduct sensitivity analysis to find the most sensitive parameters to each observable. In cases when an observable is insensitive to a parameter, even if the parameter is non-identifiable, one can still use the model to calibrate that observable.
V. Calibrate the most sensitive and identifiable model parameters. Estimate the model compartments and the reproduction number.
VI. Check model robustness assuming different types of noise in the data. Justify model predictions with the support of sensitivity analysis results. If the model is robust to noise, then one can proceed to (VII), otherwise one should go back to (II) and fix some model parameters or employ other models.
VII. Predict the future development of the epidemic with uncertainties assuming the current control measures. Then investigate how policy changes could influence the transmission dynamics of the epidemic.

We evaluate the effectiveness of the proposed framework by applying all the outlined steps to the outbreak dataset in NYC. One of the advantages of the framework’s generality is that it is not limited to a single dataset or model. In general, it provides a guideline on how to build an effective and trustworthy epidemiological model with the available data. For illustration purposes, we show how our framework can handle different data types by assuming scenarios when some observables in the NYC dataset are missing. However, for practical use, one should determine the data that are fed to the model at the very beginning.

## (I) Data acquisition and event timeline

We use the data consisting of daily cases, hospitalizations, and deaths between February 29, 2020, and February 4, 2021, to fit the model’s parameters. All the data used in this paper were extracted from the NYC’s government’s website and collected by the NYC Health Department [25]. Hospitalization data were collected from several sources, such as NYC public hospitals, non-public hospitals, and the Health Department’s syndromic surveillance database, which track hospital admissions across NYC. The data on the NYC online repository [25] is published by the date of the event, rather than the date of the report. A person is classified as a confirmed COVID-19 case when they test positive in a molecular test (PCR). A deceased individual is classified as a disease-related death if they were an NYC resident who had a positive PCR test for the virus within the last 60 days.

Since the first case of COVID-19 was reported in NYC on February 29, 2020, local authorities have implemented several non-pharmaceutical interventions, such as social distancing and mask-wearing [24, 25]. These restrictions were later relaxed as the incidence decreased. Any control measures implemented will have an impact on the transmission term *β*. Thus, we identified seven time periods defined based on the interventions implemented (also see Fig 2):

**Fig 2.**
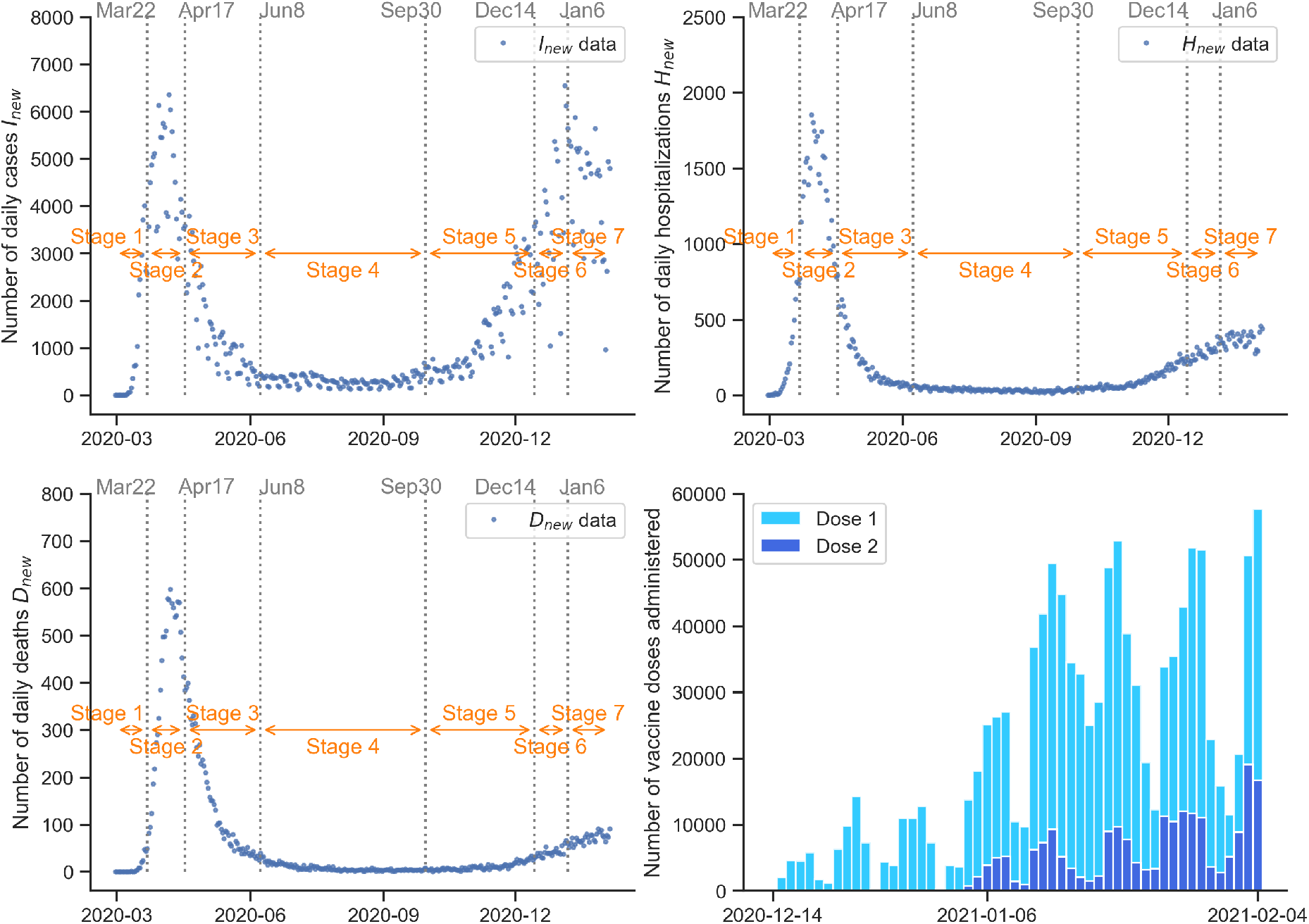
COVID-19 epidemic in New York City: data and event timeline. (a) Daily infectious population (February 29, 2020 – February 4, 2021). We split the data into seven time periods based on interventions implemented. The starting times of interventions are shown on the top of each subfigure. (b) Daily hospitalized population (February 29, 2020 – February 4, 2021). (c) Daily deceased population. (February 29, 2020 – February 4, 2021). (d) Daily vaccinated population. (December 14, 2020 – February 4, 2021).

- **Stage 1 (no control): February 29, 2020 – March 22, 2020** The New York State governor declared a state of emergency on March 7, 2020, after 89 positive cases were identified [27]. However, most businesses operated as usual until March 14, 2020, when some public libraries closed. Nightclubs, theaters, and concert venues followed suit on March 17, 2020.
- **Stage 2 (stay-at-home order): March 22, 2020 – April 17, 2020** A stay-at-home order (also known as PAUSE) issued by the New York State’s governor’s office went into effect on March 22, 2020. The PAUSE plan comprised a 10-point policy that mandated all non-essential businesses statewide to close, cancel gatherings of any size, and businesses that provide essential services must facilitate social distancing of at least six feet, among others. Schools and universities closed and moved to remote instruction.
- **Stage 3 (mask mandate): April 17, 2020 – June 8, 2020** New York State was one of the first states to issue orders mandating face coverings in public spaces [28]. This decision followed the Centers for Disease Control and Prevention (CDC) guidelines, which encouraged people to wear masks to prevent transmission of the virus through droplets generated when an infected person coughs or sneezes [29].
- **Stage 4 (four-phase reopening): June 8, 2020 – September 30, 2020** The stay-at-home order was effective in bringing down the disease’s incidence. As a result, a four-phase reopening plan was developed, taking into account seven health metrics the city needed to meet before reopening [30]. NYC entered Phase 1 of reopening on June 8, Phase 2 on June 22, Phase 3 on July 6, and Phase 4 on July 20. Each phase had specific policies that determined what businesses could reopen and in what capacity. Industries that posed the lowest risk of infection for employees and customers were allowed to reopen in Phase 1. These included but were not limited to construction, manufacturing, and wholesale supply-chain businesses and retailers for curbside pickup, in-store pickup, or drop-off [31]. Phase 2 allowed offices, places of worship (25% capacity), finance and insurance, administrative support, among others, to reopen if they follow established social distancing guidelines [32]. In Phase 3, some personal services were allowed to reopen; indoor dining was not allowed in NYC, even though this phase allowed other areas of the state to enable indoor dining at a reduced capacity. NYC entered reopening Phase 4 on July 20. Restrictions regarding group gatherings were eased, and meetings of up to 50 people were allowed. Indoor religious meetings were allowed to resume at 33% capacity. Malls, zoos, and botanical gardens were also permitted to reopen in this phase. NYC stayed at Phase 4 until September 30, 2020, when indoor dining at 25% capacity was allowed.
- **Stage 5 (indoor dining reopens at reduced capacity): September 30, 2020 – December 14, 2020** The careful reopening process, which followed the strict stay-at-home order, maintained a low infection rate in the city (below 1%) [33]. NYC resumed indoor dining services at 25% capacity on September 30, 2020, intending to double the capacity if infection rates remained low. Restaurants were required to follow an extensive set of rules upon reopening, such as temperature checks, contact tracing reporting, mask usage except when seated, and a midnight curfew.
- **Stage 6 (indoor dining closes and vaccination begins): December 14, 2020 – January 6, 2021** In December 2020, the increasing rate of virus transmission in NYC threatened to overwhelm hospital capacity. Although contact tracing data from NYC placed indoor dining as the fifth source of new infections in the state, the CDC designated indoor dining as a “high risk” activity [34]. The governor’s decision to ban indoor dining was an attempt to halt the steep increase in cases and avoid a broader shutdown. In the same week when the governor’s office closed indoor dining again, the first coronavirus vaccine was administered in Queens on December 14, 2020 [35].
- **Stage 7 (Christmas and New Year holiday ends): January 6, 2021 – February 14, 2021** The increased social activities during Christmas and New Year celebrations had a significant impact on the outbreak’s spread. This final period is defined by an event, the end of the holidays, and restaurants’ reopening in limited capacity announced for February 14, 2021. Two shipments from drug companies Pfizer and Moderna aim to cover a quarter of the estimated 1.8 million people deemed high priority to receive the vaccine in the first phase of distribution in the state. However, even though vaccination started on December 14, 2020, people vaccinated do not develop immunity to the virus immediately. The Pfizer vaccine’s first dose needs about 14 days to be 52% effective. The second dose should be administered three weeks after the first dose. The reported effectiveness of the Pfizer vaccine is 95%, while Moderna reports 94.1% [36, 37], if two doses are received. It’s worth noting that if a person only receives one dose, its effectiveness varies depending on the company that produced the vaccine. For example, the Pfizer-BioNTech vaccine is roughly 52% effective after the first dose, while the Moderna vaccine can provide 80.2% protection after one dose [36]. At–risk groups, such as older people in nursing homes and public health professionals, are prioritized for vaccination in NYC in this first phase. We use data reported by NYC Health, extracted from the Citywide Immunization Registry (CIR). The data include the daily numbers of individuals who have received the first and second dose [35]; see Fig 2.

## (II) Epidemiological Model development

Classic SIR and SEIR models do not include the exposed and presymptomatic periods, which play an important role in this particular disease. Recent studies have revealed the role of asymptomatic [41–44] and presymptomatic individuals [45–47] in the disease’s transmission chain. Additionally, due to CDC recommendations, infected individuals are required to self-isolate once they test positive for ten days [29]. Therefore, these individuals are removed from the population until they are no longer infectious. We include these individuals in the isolated compartment of our model. Therefore, to account for the complete epidemiological characteristics of a COVID-19 infection, in this model we modify an SEIR model to include presymptomatic, asymptomatic, hospitalized, isolated, and deceased compartments; see Fig 3.

**Fig 3.**
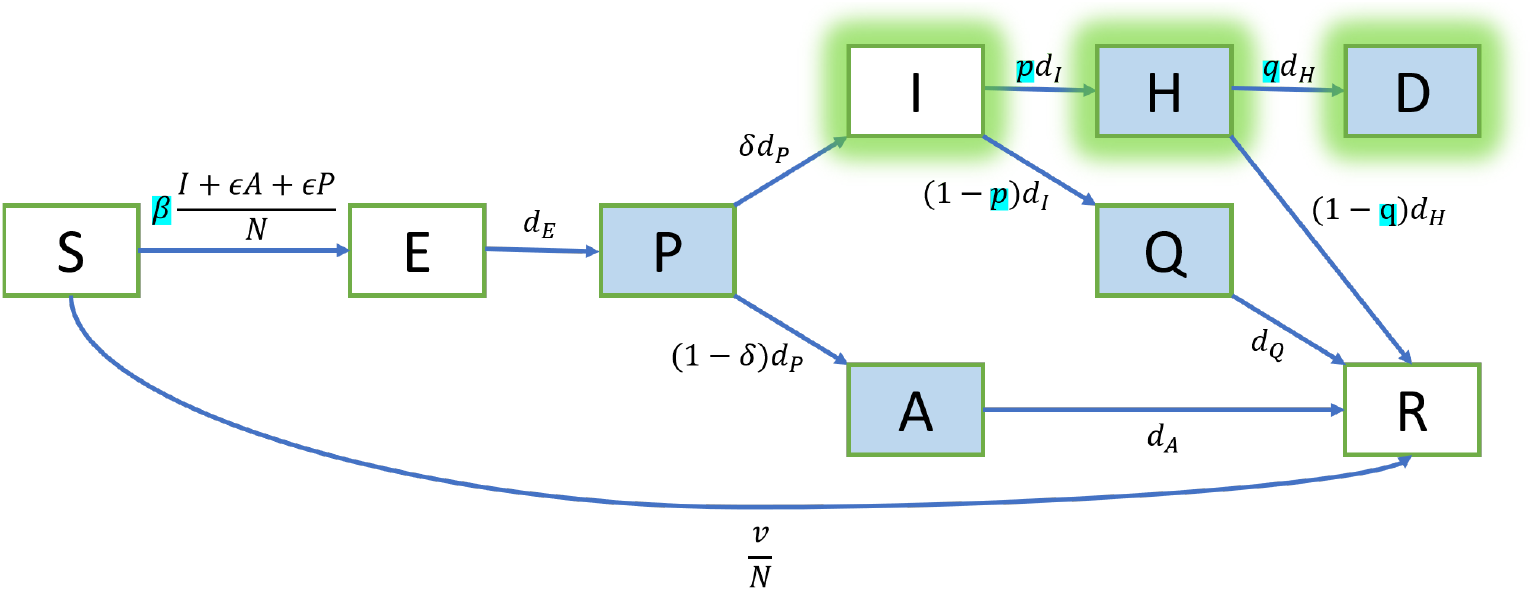
Transition diagram between epidemiological classes. We modify the classic SEIR model to include presymptomatic (*P*), asymptomatic (*A*), hospitalized (*H*), isolated (*Q*), and deceased (*D*) individuals. The given data are the inflows of symptomatic (*I*), hospitalized (*H*), and deceased (*D*) individuals. The parameters to estimate are (*β, p, q*). See Table 1 for the notations and the initial values. See Table 2 for the parameters. See Eq. (1) for the corresponding ODE system.

These modifications allow a more accurate description of the biology of the disease. Since this study focuses on a single outbreak, births and other deaths are not considered. When the epidemic begins, all individuals are susceptible and transit to the exposed class via contact with presymptomatic, symptomatic, or asymptomatic individuals. Moreover, we assume that there is no reinfection. In other words, once an individual recovers, they do not become susceptible again. We divide the total population into nine different compartments: susceptible (*S*), exposed (*E*), presymptomatic infected (*P*), symptomatic infected (*I*), asymptomatic infected (*A*), hospitalized (*H*), isolated (*Q*), deceased (*D*), and recovered (*R*). The contribution to the transmission of COVID-19 from asymptomatic individuals (*A*) relative to the transmission from symptomatic individuals (*I*) is labeled *ϵ*. Therefore, if *ϵ* = 0.75, this means that an asymptomatic individual is 75% as infectious as an asymptomatic individual. Similar to [39], we assume that presymptomatic individuals (*P*) are just as infectious as asymptomatic individuals (*A*). The transmission rate of the disease is denoted by *β*. After a latent period (1*/d*_*E*_), an exposed individual becomes presymptomatic infectious. A presymptomatic infectious individual develops symptoms with probability *δ* or is asymptomatic with probability (1 *− δ*) after (1*/d*_*P*_) days. Asymptomatic individuals recover after an infective period of (1*/d*_*A*_). A proportion *p* of symptomatic individuals is hospitalized after an infective period of (1*/d*_*I*_), while the rest of them are isolated (for example, isolated at home) but do not go to the hospital. The isolation period is (1*/d*_*Q*_). The hospitalization period is (1*/d*_*H*_), at the end of which, a proportion *q* of the hospitalized individuals die while the rest of them recover. Asymptomatic cases pose a challenge to identify since there is no widespread systematic testing of the population [41]. Thus, there is a wide range of estimates for the proportion of symptomatic individuals *δ*. In this study, we use the best estimate of *δ* provided by the CDC [38].

**Table 1.** Notations and initial values for the model in Fig 3.

| Notation | Meaning | Initial value | Note |
| --- | --- | --- | --- |
| $N$ | Total population | 8399000 | We assume constant total population in NYC |
| $S$ | Susceptible | 8398713 | $S(0) = N - E(0) - P(0) - I(0) - A(0) - H(0) - Q(0) - D(0) - R(0)$ |
| $E$ | Exposed | 270 | $E(0) = E_*/\delta$, where $E_*$ is the number of cases from day $(2/d_P + 1)$ to day $(2/d_P + 2/d_E)$ (Mar 6-10, 2020) |
| $P$ | Presymptomatic | 15 | $P(0) = P_*/\delta$, where $P_*$ is the number of cases from day 1 to day $2/d_P$ (Mar 1-5, 2020) |
| $I$ | Symptomatic | 1 | $I(0) = 1$ from data |
| $A$ | Asymptomatic | 1 | $A(0) = I(0) \times (1 - \delta)/\delta \approx 1$ |
| $H$ | Hospitalized | 0 | $H(0) = 0$ from data |
| $Q$ | Isolated | 0 | $Q(0) = 0$ |
| $D$ | Deceased | 0 | $D(0) = 0$ from data |
| $R$ | Recovered | 0 | $R(0) = 0$ |
| $I_{sum}$ | Cumulative cases | 1 | $I_{sum}(0) = 1$ from data |
| $H_{sum}$ | Cumulative hospitalizations | 0 | $H_{sum}(0) = 0$ from data |
| $D_{sum}$ | Cumulative deaths | 0 | $D_{sum}(0) = 0$ from data |

**Table 2.** Parameters for the model in Fig 3.

| Parameter | Meaning | Calibration | Range/Value | Note |
| --- | --- | --- | --- | --- |
| $\beta$ | Transmission rate | fitted | $[0, 1]$ | Fitted within wide range |
| $p$ | Hospitalization ratio | fitted | $[0, 1]$ | By definition |
| $q$ | Death from hospital ratio | fitted | $[0, 1]$ | By definition |
| $\epsilon$ | Infectivity ratio of asymptomatic to symptomatic | fixed | 0.75 | Ref. [38] |
| $\delta$ | Proportion of symptomatic individuals | fixed | 0.6 | Ref. [38] |
| $1/d_E$ | Latent period | fixed | 2.9 days | Ref. [39] |
| $1/d_P$ | Mean infectious period of $P$ class | fixed | 2.3 days | Ref. [39] |
| $1/d_I$ | Mean infectious period of $I$ class | fixed | 2.9 days | Ref. [39] |
| $1/d_A$ | Mean infectious period of $A$ class | fixed | 7 days | Ref. [40] |
| $1/d_H$ | Mean duration of $H$ class | fixed | 6.9 days | Ref. [38] |
| $1/d_Q$ | Mean duration of $Q$ class | fixed | 10 days | Ref. [29] |

Moreover, the proposed model has considered the vaccination deployment in NYC and the impact on the city’s daily cases, hospitalizations, and deaths. A susceptible individual leaves the (*S*) class and joins the recovered individuals (*ℛ*) once they are effectively vaccinated, i.e., they are vaccinated and the vaccine prevents them from getting the disease in the future. To calculate the number of effectively vaccinated individuals that takes into account the vaccine efficacy, we use a weighted sum of the number of the first doses and second doses administered. See Fig 3, where *v* is defined as the daily number of effectively vaccinated individuals.

The ODE system for our model (see Fig 3) is the following:

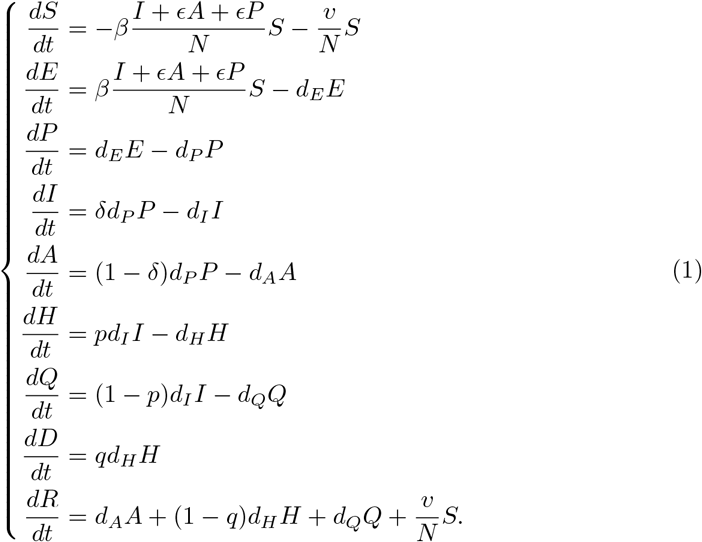

Note that the compartments (*I, H, D*) represent the current symptomatic individuals, hospitalizations, and deaths. In order to represent the daily cases *I*_*new*_, hospitalizations *H*_*new*_, and deaths *D*_*new*_, we add the following ODEs that take into account the inflow of the these compartments to record the cumulative cases *I*_*sum*_, hospitalizations *H*_*sum*_, and deaths *D*_*sum*_:

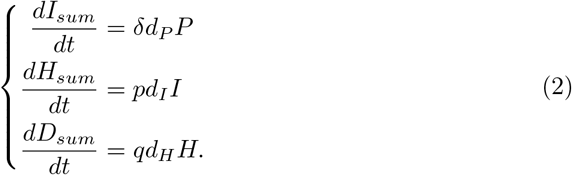

Now, the daily numbers are just the increments of the cumulative numbers:

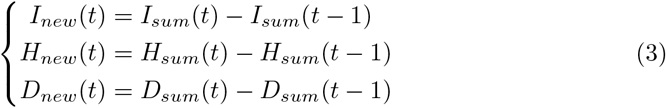

for *t* = 1, 2, 3, ….

### Time-dependent model parameters (piecewise constant *β, p*, and *q*)

The transmission rate of a disease, *β*, is the *per capita* rate of infection when a contact occurs. Directly measuring the transmission rate is not possible for most infections [48]. Nevertheless, if we want to quantify the effects of public health policies that directly impact the transmission rate, estimating this value accurately is critical. Moreover, public health policy and the discovery of better therapies and treatments affect other parameters besides the disease’s transmission rate. Notably, the percentage of disease-related deaths changes over the course of the outbreak [49]. Similarly, the hospitalization ratio varies due to increased resources channeled to the healthcare system in the city [25, 50]. Control measures implemented in NYC and the subsequent relaxation of restrictions impact the incidence curve in different ways—most of them non-linear. Therefore, defining the transmission rate *β*(*t*) as a piecewise constant function is a simplification that allows us to estimate the impact each policy has in each stage. Similarly, we define the piecewise constant hospitalization ratio *p*(*t*) and death from hospital ratio *q*(*t*), which also exhibit varying values over time:

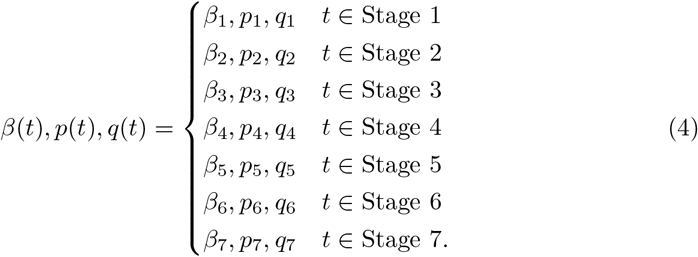

We fit the parameters in each stage defined by policy changes, such as the stay-at-home order and the subsequent reopening processes; see Fig 2.

### Reproduction number

The basic (control) reproduction number, denoted by *ℛ*_0_ (*ℛ*_*c*_), is the average number of secondary infections caused by one infected individual in an entirely susceptible well-mixed population in the absence (presence) of disease control. The control reproduction number of the model is:

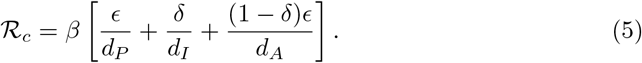

In this particular study, given that the model parameters are defined in a piecewise fashion and there was no control in Stage 1, the basic reproduction number, *ℛ*_0_, is computed by using *β* = *β*_1_.

The transmission of the disease slows down when there are more immune individuals.Since *ℛ*_*c*_ is the number in an entirely susceptible population, we can calculate the effective reproduction number:

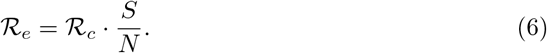

By setting *ℛ*_*e*_ = 1, we obtain the immunity threshold of the ODE system, which is the critical portion of the population needed to be immune to stop the transmission of the disease:

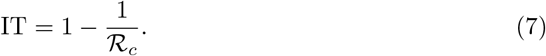

The herd immunity threshold (HIT) is calculated by substituting *ℛ*_*c*_ with *ℛ*_0_. A higher *ℛ*_0_ results in a higher HIT.

## (III) Identifiability analysis

In this section we address whether a set of unknown parameters in the proposed model is globally identifiable from the available data. Fitting a model to the data is not sufficient to show how reliable the estimated parameters are. Insufficient or noisy data can produce drastically different sets of parameters without affecting the fit to data if a model is non-identifiable [2]. Furthermore, depending on the available data (observables), different models may be appropriate.

Formally speaking, a parameter in a dynamical system is considered to be identifiable if the solutions can uniquely determine it. Two different types of identifiability, namely *structural* and *practical* identifiability are considered in this paper. Structural identifiability analysis studies the uniqueness of parameter values from the perspective of the structure of the equation and is normally conducted before the fitting of the model, thus commonly referred to as a priori identifiability. Global (structural) identifiability provides conclusions about a parameter’s identifiability in the entire parameter space [51–54]. In particular, it guarantees the opportunity of uniquely identifying the model parameters from the data [51–53, 55]. In some cases, however, *local* structural identifiability may be sufficient, and hence the range of values of the parameter to be identified should be limited. On the other hand, practical identifiability analysis mainly addresses the issue of nonuniqueness when fitting the model on the discrete data points, i.e., a posteriori. Structural identifiability does not imply practical identifiability because of the amount and quality of the data. A detailed explanation of these two types of identifiability can be found in S4 Text. We use the open-source software SIAN [56] for structural identifiability and use correlation matrix calculated from Fisher Information Matrix (FIM) for practical identifiability. Details of the implementation can also be found in the supporting information.

In our framework, we analyze both structural and practical identifiability, and use the results as guidelines for parameter selection. The importance of performing both types of analysis resides in the fact that structural identifiability itself does not guarantee the goodness-of-fit of the model. It turns out that in our case, fitting all structural identifiable parameters would lead to practically non-identifiable results.

### Structural identifiability

There are 11 undetermined parameters in the proposed model, and it is impossible to fit every parameter without fixing some of the values. For example, it is unnecessary to fit biologically determined parameters such as the time an individual spends in the exposed or infected classes. Since *d*_*E*_, *d*_*P*_, *d*_*I*_, *d*_*A*_, *d*_*H*_, and *d*_*Q*_ are determined by the biology of the disease, we fix these values according to [39].

We analyze the structural identifiability of the rest of the parameters when different types of data are given. Note that for general use of our modeling framework, one should fix the dataset at step (I). Here we consider all the scenarios just for illustration purpose. Specifically, we assume that the data are given as the cumulative cases *I*_*sum*_, cumulative hospitalizations *H*_*sum*_, and cumulative deaths *D*_*sum*_, or a subset of the three aforementioned observables, because these quantities can be calculated directly from daily quantities *I*_*new*_, *H*_*new*_, and *D*_*new*_. In other words, it is equivalent to assume *X*_*new*_ or *X*_*sum*_ to be given as one of the observables, where *X* can be *I, H*, and *D*. The effectively vaccinated population *v* is treated as the input variable to the system.

According to Table 3, when only two out of three observables are available, the model is not identifiable and the fitting result will not be unique. The results are to be interpreted in the following way. In the case of lacking deceased individual counts, the death rate *q* cannot be inferred accurately. If the hospitalization data are not available, both the hospitalization ratio *p* and death from hospital ratio *q* cannot be inferred accurately. When the infectious data are missing, results for the transmission rate *β* and the hospitalization ratio *p* cannot be inferred accurately. Non-identifiability of parameters does not imply the failure of the model. In (VI), we demonstrate that our model can still be robust and predictable in some cases when it is not identifiable since the non-identifiable parameters could be insensitive.

**Table 3.** Structural identifiability of *β, p, q, ϵ, δ* with different observables. Global/not means structurally globally/not identifiable, respectively. We fix all the rest of the parameters as in Table 2. It is necessary to include all three observables to guarantee the structural identifiability of the model.

| Parameter | $I_{sum}, H_{sum}, D_{sum}$ | $I_{sum}, H_{sum}$ | $I_{sum}, D_{sum}$ | $H_{sum}, D_{sum}$ | $I_{sum}$ | $H_{sum}$ | $D_{sum}$ |
| --- | --- | --- | --- | --- | --- | --- | --- |
| $\beta$ | global | global | global | not | global | not | not |
| $p$ | global | global | not | not | not | not | not |
| $q$ | global | not | not | global | not | not | not |
| $\epsilon$ | global | global | global | global | global | global | global |
| $\delta$ | global | global | global | global | global | global | global |

The analysis above shows that it would be hard to draw conclusions about the fitting correctness of transmission rate, proportion of isolated individuals, and proportion of disease-related deaths when one of the observables considered in this paper is missing. One of the main differences between the proposed model and most other existing SEIR-based models is that our model integrates information of infectious, hospitalized, and deceased populations simultaneously, therefore producing more reliable results on these estimated parameter values. Since we have data for all three observables in NYC, we should utilize all of them.

In practice, hospitalization data could be reported in different ways; some databases provide daily reports of the number of hospitalized individuals, whereas others register the number of currently hospitalized individuals. Regardless of the data type available, structurally identifiability of the model remains the same according to S2 Table.

### Practical identifiability

We then proceed with fitting 5 undetermined parameters using all the available data, i.e., *I*_*sum*_, *H*_*sum*_, and *D*_*sum*_. The model-fitting techniques, including the loss function and optimization method, are detailed in the next section. The fitted parameter values can be found in Table 4. We see that the values of *ϵ* and *δ* vary a lot among different stages, which is inconsistent with reality. This poses a question on the practical identifiability of the model.

**Table 4.** Practical identifiability and estimation of parameters when fixing. *d*_*E*_, *d*_*P*_, *d*_*I*_, *d*_*A*_, *d*_*H*_, *d*_*Q*_. The symbol ✓/ ✗ means practically identifiable/not identifiable, respectively. The fitted values will **not** be counted towards our final result because the model is not identifiable in this case.

| Parameter | Identifiable | Stage 1 | Stage 2 | Stage 3 | Stage 4 | Stage 5 | Stage 6 | Stage 7 |
| --- | --- | --- | --- | --- | --- | --- | --- | --- |
| $\beta$ | × | 0.64 | 0.18 | 0.32 | 0.18 | 0.37 | 0.19 | 0.48 |
| $p$ | ✓ | 0.39 | 0.31 | 0.15 | 0.12 | 0.08 | 0.08 | 0.08 |
| $q$ | ✓ | 0.14 | 0.37 | 0.35 | 0.22 | 0.13 | 0.16 | 0.20 |
| $\epsilon$ | × | 1.00 | 1.00 | 0.05 | 0.80 | 0.34 | 1.00 | 0.00 |
| $\delta$ | × | 1.00 | 0.95 | 0.69 | 0.63 | 0.70 | 0.58 | 0.71 |

The correlation matrices are then computed based on parameters obtained in Stage 1 to help determine the practical identifiability. In practice, one can obtain the correlation matrix from the FIM of model parameters, with details given in S4 Text. As shown in Fig 4(b), there is a strong correlation between *ϵ, δ*, and *β*, while either *p* or *q* is uncorrelated with the rest of the parameters. The same phenomenon is observed when we calculate the correlation matrix based on parameters obtained in other stages; see S2 Fig. This indicates that *ϵ, δ*, and *β* are not practically identifiable. Two of them need to be fixed, while the rest and *p, q* can be fitted. In this paper, we fix *ϵ, δ* and fit *β, p, q* for the following reason: *β*, which represents the transmission rate, is highly affected by local government policy and does not have a stable and universal value compared to *ϵ* and *δ*. This means that the value of *β* could be very different across different datasets and is hard to determine a priori. Therefore, we fix *ϵ* and *δ* according to [38]. After that, the model becomes practically identifiable as shown in S3 Fig. A summary of the reasoning is shown in Fig 4(a).

**Fig 4.**
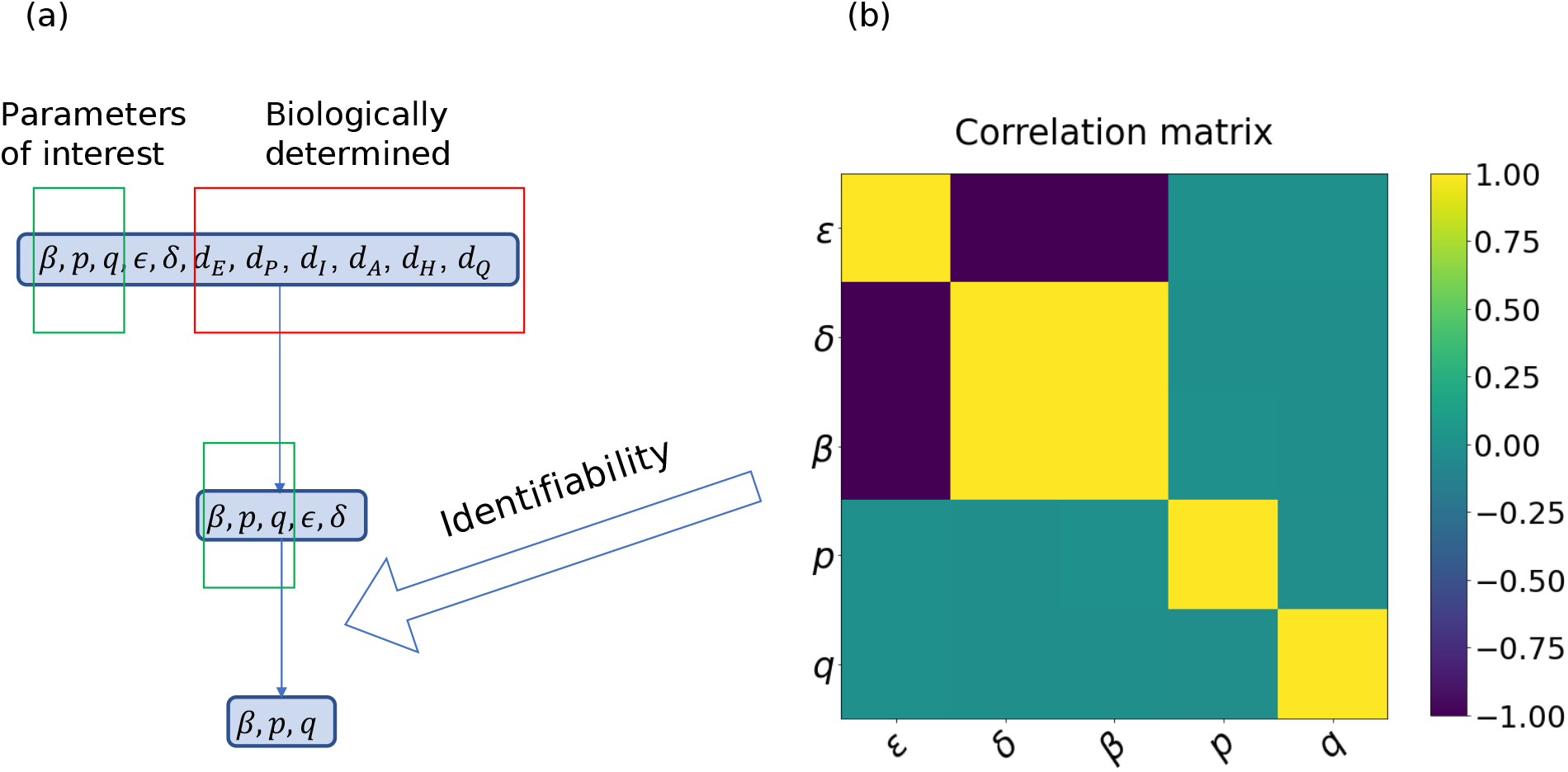
The procedure of choosing parameters to fit. (a) The procedure of determining parameters to fit. We fix *d*_*E*_, *d*_*P*_, *d*_*I*_, *d*_*A*_, *d*_*H*_, *d*_*Q*_ because they are biologically determined, and then fix *ϵ, δ* due to the result from the correlation matrix analysis. (b) The correlation matrix of five parameters. Each colored off-diagonal cell represents the correlation between two parameters. Green means (almost) not statistically correlated while yellow/purple represents positively/negatively correlated, respectively.

## (IV) Sensitivity analysis

Variance-based sensitivity analysis, also called Sobol sensitivity analysis, is a global method that measures sensitivity across the whole input space. It decomposes the model’s output variance into fractions that can be attributed to individual inputs or groups of inputs [20]. Suppose we are given a black box model:

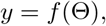

where *y* ∈ ℝ is the output and Θ = [*θ*_1_, *θ*_2_, …, *θ*_*k*_] ∈ [0, 1]^*k*^ are independent and uniformly distributed uncertain inputs. If some components of Θ are not within [0, 1], we may transform Θ into the unit hypercube.

First-order sensitivity index measures the contribution to the output variance by a single input *θ*_*i*_ alone:

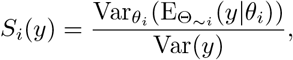

where Θ_*∼i*_ = [*θ*_1_, …, *θ*_*i−*1_, *θ*_*i*+1_, …, θ_*k*_]. Total-order sensitivity index measures the contribution to the output variance by an input, including its first-order effect and all higher-order interactions with other inputs:

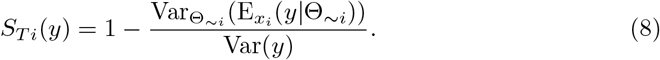

Note that

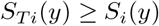

by definition, and

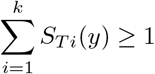

since the interaction between *θ*_*i*_ and *θ*_*j*_ is counted in both *S*_*T i*_(*y*) and *S*_*T j*_(*y*).

Sensitivity analysis does not rely on any data. Instead, it analyzes the dependence relationship between the outputs and the inputs of a given model from the level of parametric equations when a specific initial condition to the system is given. Regarding the model in Fig 3, the cumulative infectious population *I*_*sum*_ in each stage of the pandemic is a function of the parameters *β, p*, and *q* (they are assumed constant during each period). So are the cumulative hospitalized population *H*_*sum*_ and the cumulative death population *D*_*sum*_. Using Sobol’s method, we obtain the first-order sensitivity and total-order sensitivity of each model output of interest (*I*_*sum*_, *H*_*sum*_, *D*_*sum*_) with respect to each parameter (*β, p, q*). The ranges for *β, p*, and *q* are [0, 1]. For each model output, 8000 samples are generated using Saltelli’s sampling scheme. The results are plotted in Fig 5. We can see that the cumulative cases *I*_*sum*_ does not depend on *p* or *q*. For the cumulative hospitalizations *H*_*sum*_, *β* is the most important parameter while *q* does not have any impact. For the cumulative deaths *D*_*sum*_, *β* is the most influential parameter as well. The qualitative relationship between the scale of sensitivity indices for different parameters is the same across all stages.

**Fig 5.**
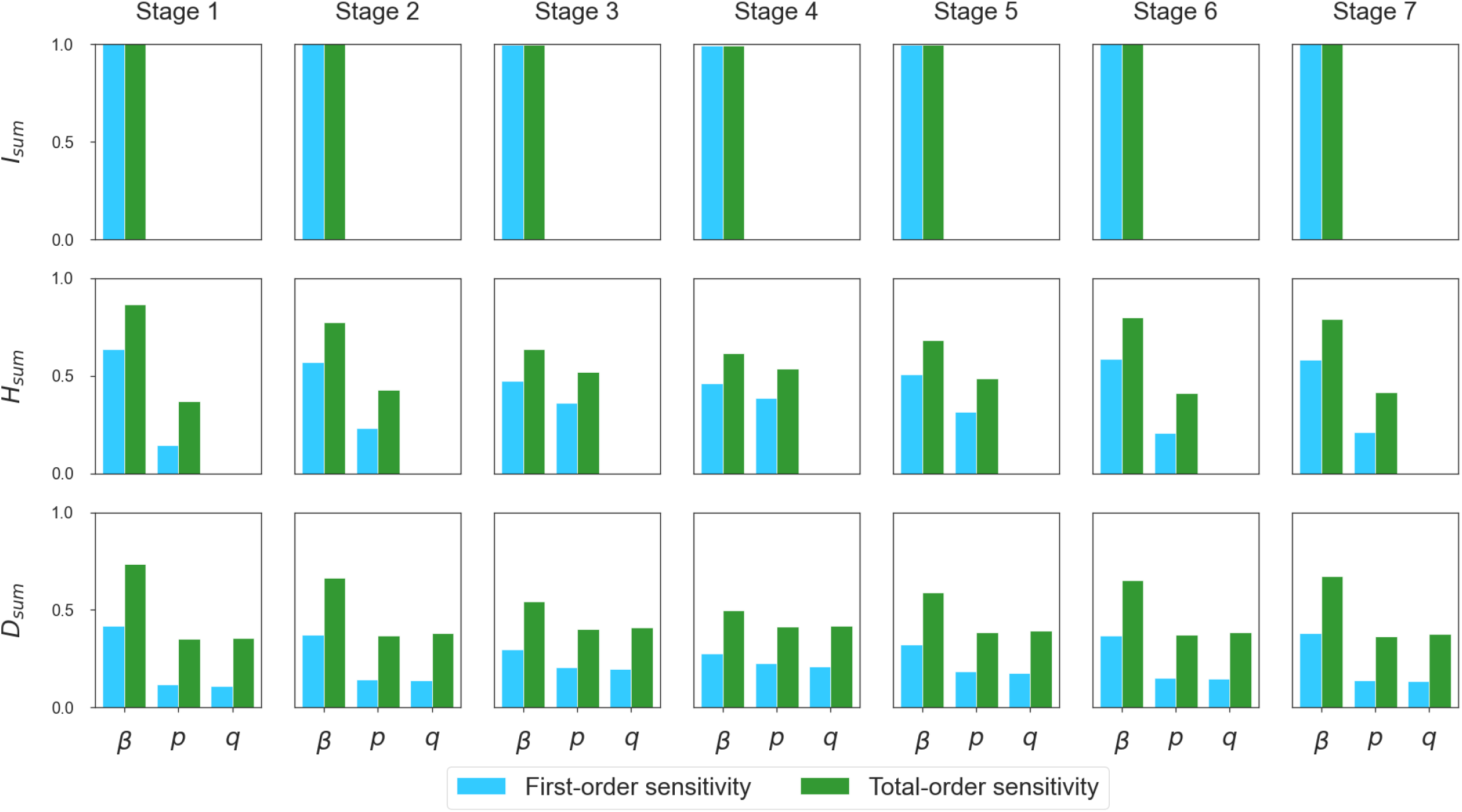
Sensitivity of each quantity of interest. (*I*_*sum*_, *H*_*sum*_, *D*_*sum*_**) with respect to each parameter (***β, p, q*). The parameter *β* is the most important parameter for all three quantities of interest in every stage of the pandemic. The parameter *p* has no influence on *I*_*sum*_. The parameter *q* has no influence on *I*_*sum*_ or *H*_*sum*_.

In the proposed model, the parameter *β* is the most important parameter for the prediction of all *I*_*sum*_, *H*_*sum*_, and *D*_*sum*_. Since *p* and *q* do not contribute to *I*_*sum*_, our model may predict *I*_*sum*_ even if *p* and *q* were inaccurate. Similarly, our model may predict *H*_*sum*_ even if *q* were inaccurate.

## (V) Model calibration

### Estimation of the number of effective vaccinations

The vaccination data of COVID-19 in NYC are given in the form of the number of first and second doses administered from December 14, 2020 to February 4, 2021 (see Fig 2). The vaccine efficacy is 52% for only one dose and 95% for both doses. As the vaccines are not 100% effective, in order to calculate the number of effectively vaccinated individuals that takes into account the vaccine efficacy, we use a weighted sum of the number of the first doses and second doses administered:

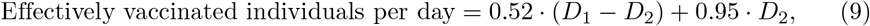

where *D*_1_ is the number of individuals who receive the first dose and *D*_2_ is the number of individuals who receive the second dose on that day. As the vaccines provide immunity 14 days after they are received, we remove the effectively vaccinated individuals from the susceptible (*S*) class and join them to the recovered (*R*) class 14 days after they have received the vaccines. To simplify the study, we approximate and predict the number of daily effective vaccinations linearly with a cap of 17, 500 per day, which corresponds to a maximum capacity of about 35, 000 total doses per day; see Fig 6. This approximated and predicted number is used as the time-dependent parameter *v* in our model (see Fig 3). Before any vaccine is effective, we have *v =* 0.

**Fig 6.**
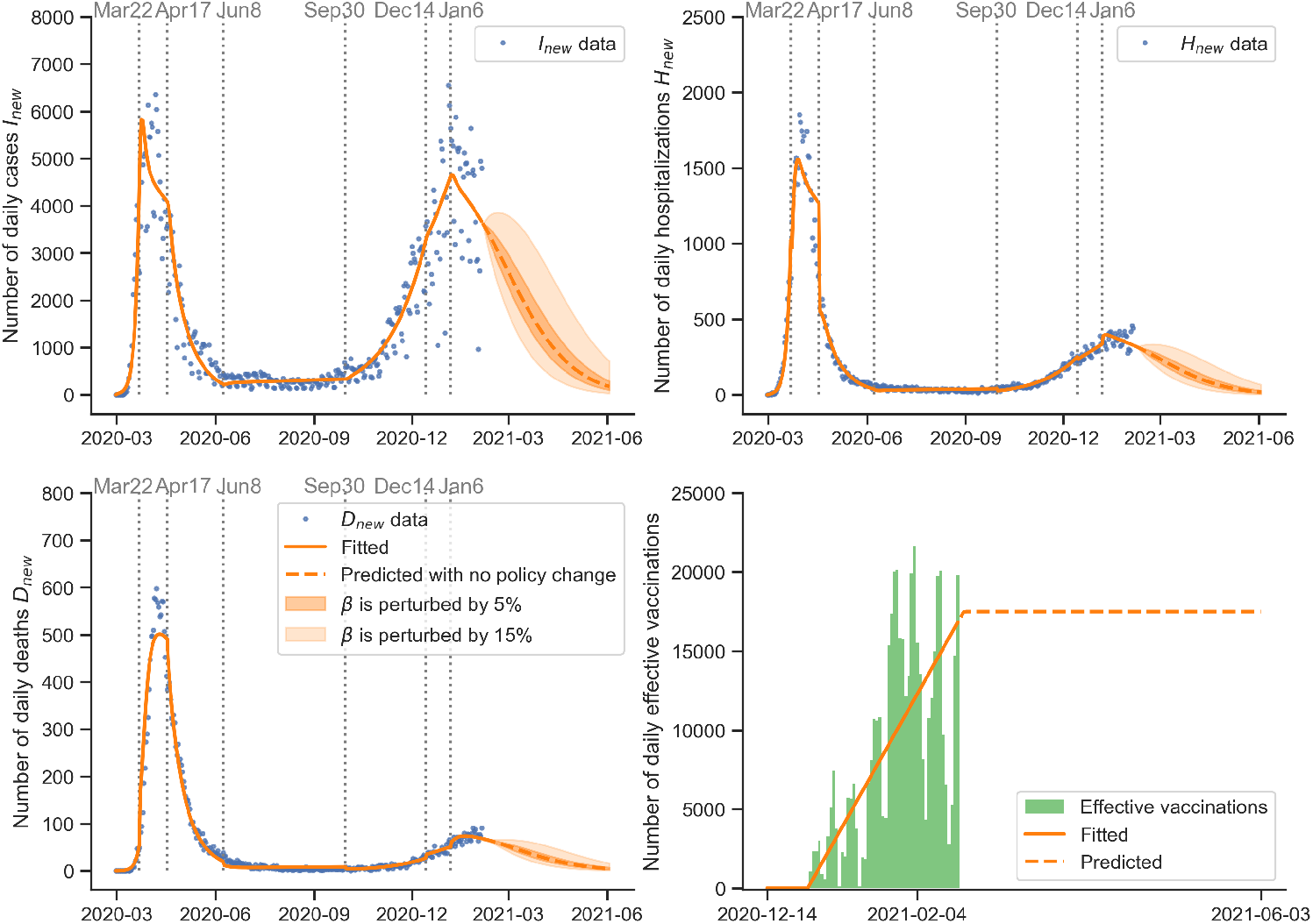
Estimation of daily cases, hospitalizations, deaths, and vaccinations in New York City. (a) Estimation of daily cases. (b) Estimation of daily hospitalizations. (c) Estimation of daily deaths. (d) We calculate the number of effective vaccinations as a weighted sum of the number of first and second doses administered as shown in Fig 2; we approximate the daily number of effective vaccinations linearly and assume it grows linearly until it reaches the maximum capacity of 17,500 per day.

### Parameter estimation via simulated annealing

The data of daily cases, hospitalizations, and deaths of COVID-19 in NYC from February 29, 2020 to February 4, 2021 are given in Fig 2. We assume a constant population size of 8.399 million people in NYC and do not consider migration. Using the data with the model in Fig 3, initial values in Table 1, and parameters in Table 2, we fit the transmission rate *β*, hospitalization ratio *p*, and death from hospital ratio *q* within the range [0, 1]. The fitting is split into seven stages defined by the public policies described in Fig 2. In each stage, the parameters (*β, p, q*) are assumed to be constant as in Eq. (4). We use simulated annealing, a global optimization algorithm, to search for the optimal parameter values in each stage. The objective is to minimize the following loss function:

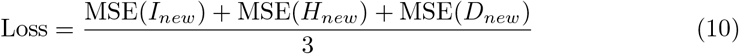

in the region *β, p, q* ∈ [0, 1], where “MSE” stands for mean squared error. The estimated final value in the previous stage is the initial value for the next stage. The results of all compartments are plotted in Fig 6 and Fig 7. The time-dependent parameters (*β, p, q*) and the control reproduction number *ℛ*_*c*_ are shown in Fig 8, Table 5, and Table 6.

**Table 5.** Estimation of parameters, control reproduction number, and immunity threshold. The transmission rate *β* and the control reproduction number ℛ_*c*_ change between different stages, indicating that local government policies in New York City and public holidays have a strong impact on the transmission dynamics of the pandemic.

|  | Meaning | Stage 1 | Stage 2 | Stage 3 | Stage 4 | Stage 5 | Stage 6 | Stage 7 |
| --- | --- | --- | --- | --- | --- | --- | --- | --- |
| $\beta$ | Transmission rate | 0.82 | 0.17 | 0.11 | 0.19 | 0.24 | 0.22 | 0.19 |
| $p$ | Hospitalization ratio | 0.41 | 0.30 | 0.13 | 0.12 | 0.08 | 0.08 | 0.09 |
| $q$ | Death from hospital ratio | 0.15 | 0.37 | 0.35 | 0.22 | 0.13 | 0.17 | 0.20 |
| $\mathcal{R}_c$ | Control reproduction number | 4.55 | 0.97 | 0.60 | 1.07 | 1.35 | 1.24 | 1.06 |
| IT | Immunity threshold | 78.0% | 0.0% | 0.0% | 6.4% | 26.2% | 19.3% | 5.6% |

**Table 6.** Percentage changes of parameters and control reproduction number between contiguous stages. The stay-at-home order in Stage 2, mask mandate in Stage 3, closing of indoor dining and starting of vaccination in Stage 6, and end of the holidays in Stage 7 lead to decreases in the transmission rate *β* and the reproduction number *ℛ*_*c*_. The four-phase reopening in Stage 4 and reopening of indoor dining in Stage 5 lead to increases in *β* and *ℛ*_*c*_.

|  | Meaning | Stage 1–2 | Stage 2–3 | Stage 3–4 | Stage 4–5 | Stage 5–6 | Stage 6–7 |
| --- | --- | --- | --- | --- | --- | --- | --- |
| $\beta$ | Transmission rate | −78.6% | −38.3% | +78.0% | +26.8% | −8.6% | −14.5% |
| $p$ | Hospitalization ratio | −26.2% | −55.9% | −10.3% | −31.1% | −8.5% | +14.5% |
| $q$ | Death from hospital ratio | +150.5% | −5.3% | −38.0% | −40.7% | +28.1% | +18.9% |
| $\mathcal{R}_c$ | Control reproduction number | −78.6% | −38.3% | +78.0% | +26.8% | −8.6% | −14.5% |

**Fig 7.**
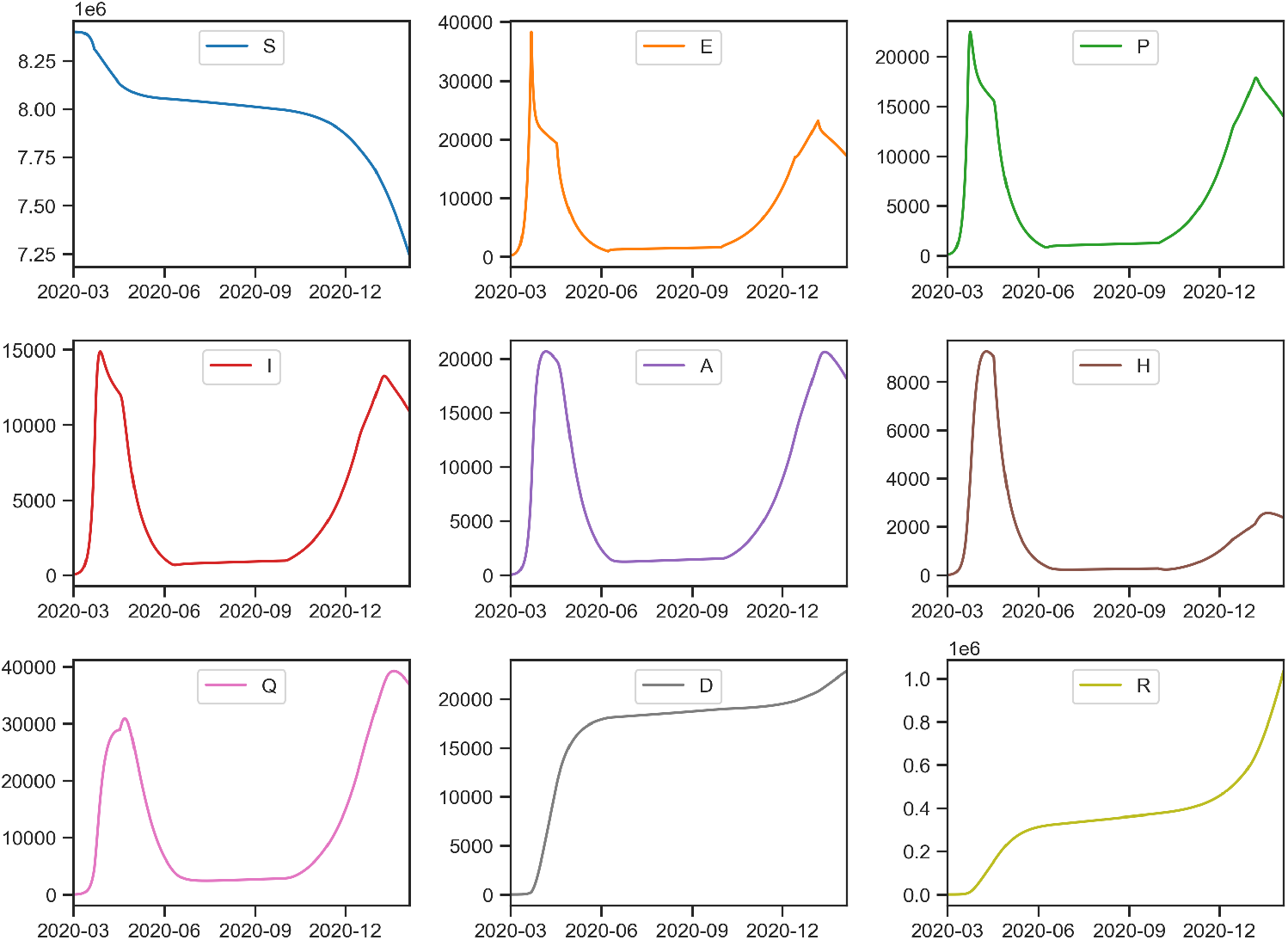
Estimation of the unobserved dynamics in all the model compartments (*S, E, P, I, A, H, Q, D, R*). The number of susceptible individuals (*S*) drops significantly as the number of cases hikes after December 2020.

**Fig 8.**
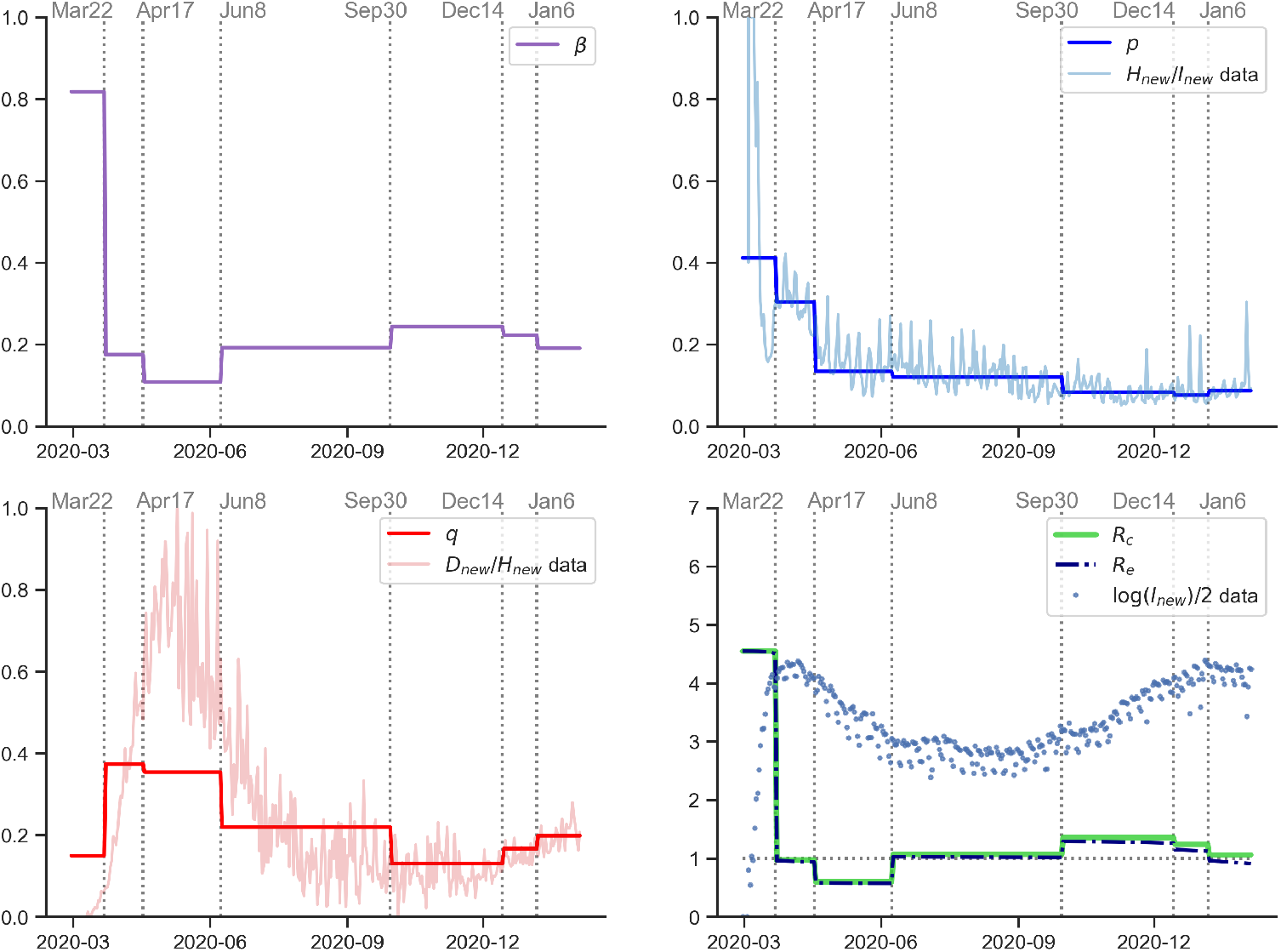
Estimation of parameters and reproduction numbers. (a) Estimated time-dependent transmission rate *β*(*t*). (b) Estimated time-dependent hospitalization ratio *p*(*t*), compared with daily hospitalizations over daily cases calculated from the raw data. (c) Estimated time-dependent death from hospital ratio *q*(*t*), compared with daily deaths over daily hospitalizations calculated from the raw data. (d) Estimated control reproduction number *ℛ*_*c*_ and effective reproduction number *ℛ*_*e*_ calculated by the estimated parameters, compared with 1*/*2 of the logarithm of daily cases.

The control reproduction number *ℛ*_*c*_, whose expression is in Eq. (5), depends on six parameters: *β, ϵ, δ, d*_*P*_, *d*_*I*_, and *d*_*A*_. Since *ϵ, δ, d*_*P*_, *d*_*I*_, and *d*_*A*_ are fixed as in Table 2, *ℛ*_*c*_ is proportional to the transmission rate *β*; see Fig 8. We plot the evolution of *ℛ*_*c*_ over time and overlay the scaled daily cases to demonstrate how the number of daily cases and *ℛ*_*c*_ (or *β*) are related to each other. Before any closures took place on March 22, 2020, we had a high *ℛ*_*c*_ with exponential growth of daily cases. Once the strict control measures were rolled out, *ℛ*_*c*_ was considerably reduced below 1 along with a decline of daily cases. During the reopening Phases 1–4, *ℛ*_*c*_ rose to around 1 with a stabilized number of daily cases. When indoor dining was reopened on September 30, 2020, *ℛ*_*c*_ rose to above 1 with another wave of daily cases. After indoor dining was closed again on December 14, 2020, *ℛ*_*c*_ decreased. After the end of holidays, *ℛ*_*c*_ further decreased.

### Bayesian posterior simulation via MCMC

We use the loss function Eq. (10) as the negative log-likelihood of the posterior distribution of the parameters (*β, p, q*). We assume that each parameter’s prior distribution is independent and uniformly distributed in [0, 1]. Using Markov chain Monte Carlo (MCMC) simulation, we may simulate the posterior distribution of each parameter associated with our approach. As before, the simulation is done within each stage where (*β, p, q*) are assumed to be constant. In each stage, four chains of 1000 samples are drawn with 200 burn-in samples in every chain. We initialize the chains at the estimation given by simulated annealing to speed up the algorithm; see S5 Fig – S11 Fig for the posterior distributions and the sampling processes. We can see that the chains are well-mixed, which implies the convergence of the sampling. The narrow posterior distributions indicate that our numerical algorithm is robust, the quantity of data is sufficient for our approach of minimizing the loss function (10), and the parameters are indeed close to constant in every stage. As a result, the parameter estimation is reliable.

## (VII) Model robustness analysis

Using the fitted parameter values in Table 5, we perform the Monte Carlo simulation to check the robustness of our model to perturbations, adapting ideas from [19, 57]. The major difference between their approach and ours is that we incorporate sensitivity analysis results in our method.

We first multiply the daily increase in the calibrated data (a subset of {*I*_*sum*_, *H*_*sum*_, *D*_*sum*_}) by independent and identically distributed Gaussian random noise of mean 1 and standard deviation *σ* to generate a new dataset, which looks like our original dataset with measurement error. Then, we estimate the parameters by fitting the model to the artificially generated dataset and compare the result with the parameter values obtained in Table 5. The same procedure is repeated for *M* = 1000 times, and we compute the average error between the parameter values estimated from the original and the generated datasets. We then rescale this error by a sensitivity constant, which measures the importance of different parameters on the observables of interest. The quantity we obtained is named sensitivity-based average relative error (SARE):

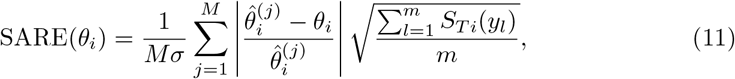

where *θ*_*i*_ is the fitted value of the *i*th parameter (i.e., *β, p, q*) on the original dataset, 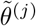 is the fitted value of the *i*th parameter on the *j*th generated dataset, and *y*_*l*_ is the *l*th observable (i.e. a subset of {*I*_*sum*_, *H*_*sum*_, *D*_*sum*_}). The quantity *S*_*T i*_(*y*_*l*_) defined in Eq. (8) measures the total contribution to the variance of *y*_*l*_ by *θ*_*i*_. In other words, it quantifies the influence of *θ*_*i*_ on the output variables. For more important parameters that have a higher *S*_*T i*_, we require more robustness.

When the parameters are piecewise constant and fitted separately, we define the overall SARE of that parameter to be the largest SARE calculated in every stage. For example, in this paper, SARE(*β*) = max_*s*∈{1,…, 7}_ SARE(*β*_*s*_). Finally, we define the maximum sensitivity-based average relative error (MSARE) of the model to be the largest SARE of all the model parameters:

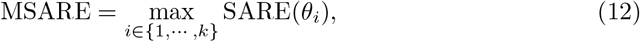

where *k* is the total number of parameters to be estimated. If MSARE *<* 1, we say the model is robust to perturbation. The algorithm is detailed in S5 Text. This method is a complement to the identifiability analysis in the sense that the identifiability analysis only takes care of the uniqueness of model parameters but not their significance to the data. An insensitive non-identifiable parameter would not influence the predictability of the model too much. On the other hand, robustness does not guarantee the correctness of the fitted parameters or the inferred compartmental values apart from the particular data. One has to combine identifiability with robustness to draw conclusions.

The first column in Fig 9 shows that when (*I*_*sum*_, *H*_*sum*_, *D*_*sum*_) are given, our model is robust to noise, which justifies the correctness of the fitting result (since *β, p, q* are also identifiable) and provides a theoretical backup for the forecasting in the next section. The other columns in Fig 9 show that even when some observables are missing, the model could still be robust to perturbation at different noise levels, so that one can use our model to make predictions in some missing observable cases. For example, only infectious and hospitalized data (*I*_*sum*_, *H*_*sum*_), only hospitalized and deceased data (*H*_*sum*_, *D*_*sum*_), only infectious data (*I*_*sum*_), or only hospitalized data (*H*_*sum*_) are available. However, in these scenarios, one should not use the model to infer other compartments without given data; see S4 Fig. Note that when *D*_*sum*_ is not available, SARE(*q*) = 0 because neither *I*_*sum*_ nor *H*_*sum*_ is sensitive to *q*. Similarly, SARE(*p*) = 0 when only *I*_*sum*_ is available since *I*_*sum*_ is not sensitive to *p*.

**Fig 9.**
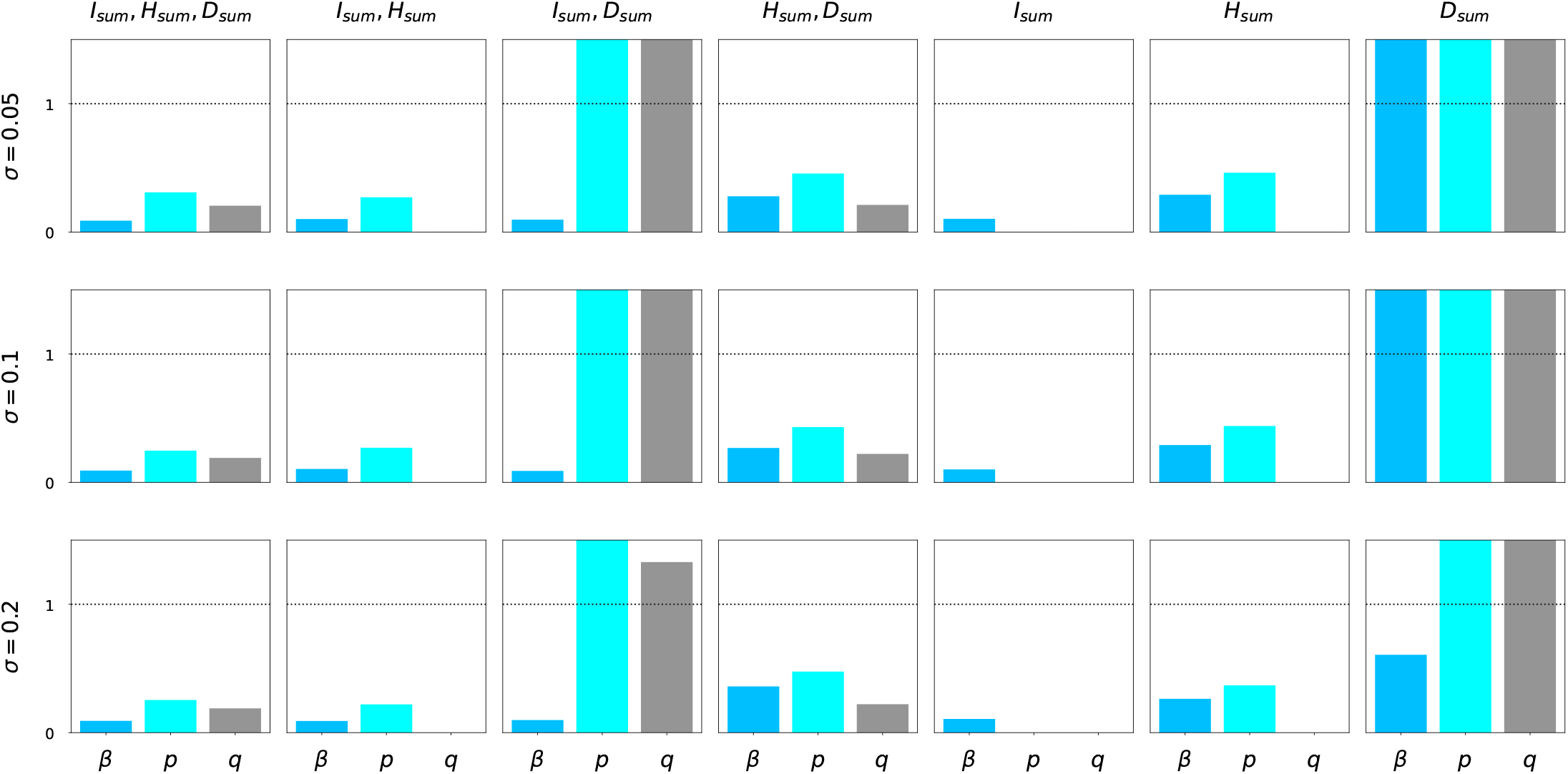
Sensitivity-based Average Relative Error (SARE) of (*β, p, q*) in different observable settings. Each row corresponds to a standard deviation level of random noise multiplied to the observables. Each column represents an observable setting. Note that when SARE of a parameter is 0, it means the provided data are not sensitive to that parameter. When (*I*_*sum*_, *H*_*sum*_, *D*_*sum*_) are given, SARE is lower than the threshold 1. Therefore, our model is robust to noise in the NYC dataset. In some of the missing observable cases, our model is also robust to perturbations at different noise levels.

## (VII) Forecasting with uncertainties and scenarios

The situation in NYC evolves day by day. The city reinstated indoor dining restrictions in mid-December due to the steady increase in the virus incidence. The ever-changing policies add a high level of uncertainty to any long term forecast we can make. Here, we explore our model’s ability to predict the number of daily cases, hospitalizations, and deaths in the city with uncertainty.

The MCMC simulation provides us with a way to quantify uncertainty. We may sample from the posterior distribution of the parameters in the last stage (see S11 Fig) and run the model after that to obtain a distribution of the predicted daily cases, hospitalizations, and deaths. However, this approach assumes that the situation remains the same after the last stage, which may not be the case. There might be policy changes or other events. As a result, we perturb the transmission rate *β* by a percentage to reflect future policy changes or other events. We sample from the posterior distribution of the parameters (*β, p, q*) in the last stage. Then, we multiply *β* by a random number drawn from a uniform distribution (𝒰 (0.95, 1.05) or 𝒰 (0.85, 1.15)). In other words, we perturb *β* by 5% or 15%. As a reference, see Table 6 for the historical changes of *β* between contiguous stages. We run the model with the initial value as the ending value of the last stage to predict daily cases, hospitalizations, and deaths. After repeating 4000 times, we obtain a distribution of the daily cases, hospitalizations, and deaths at every timestamp after the last stage. Then, the 95% confidence interval at every timestamp is plotted.

In Fig 10, we consider three different scenarios: no indoor dining, reopening indoor dining on February 14, and reopening indoor dining on March 14. The uncertainties given by the perturbed *β*’s are plotted in each scenario. For indoor dining, we assume it is a 25% reopening, which is the same as what happened in Stage 5 (September 30, 2020 to December 14, 2020). We multiply *β* by *β*_5_*/β*_6_ to represent the change of the transmission rate caused by the reopening of indoor dining. An increase in infectious, hospitalized, and deceased population is expected if the restaurants are reopened. Postponing the reopening of restaurants from February 14 to March 14 may reduce the number of infectious, hospitalized, and deceased individuals. The actual situation might vary depending on the details and implementations of the actual indoor dining policies that take place in 2021.

**Fig 10.**
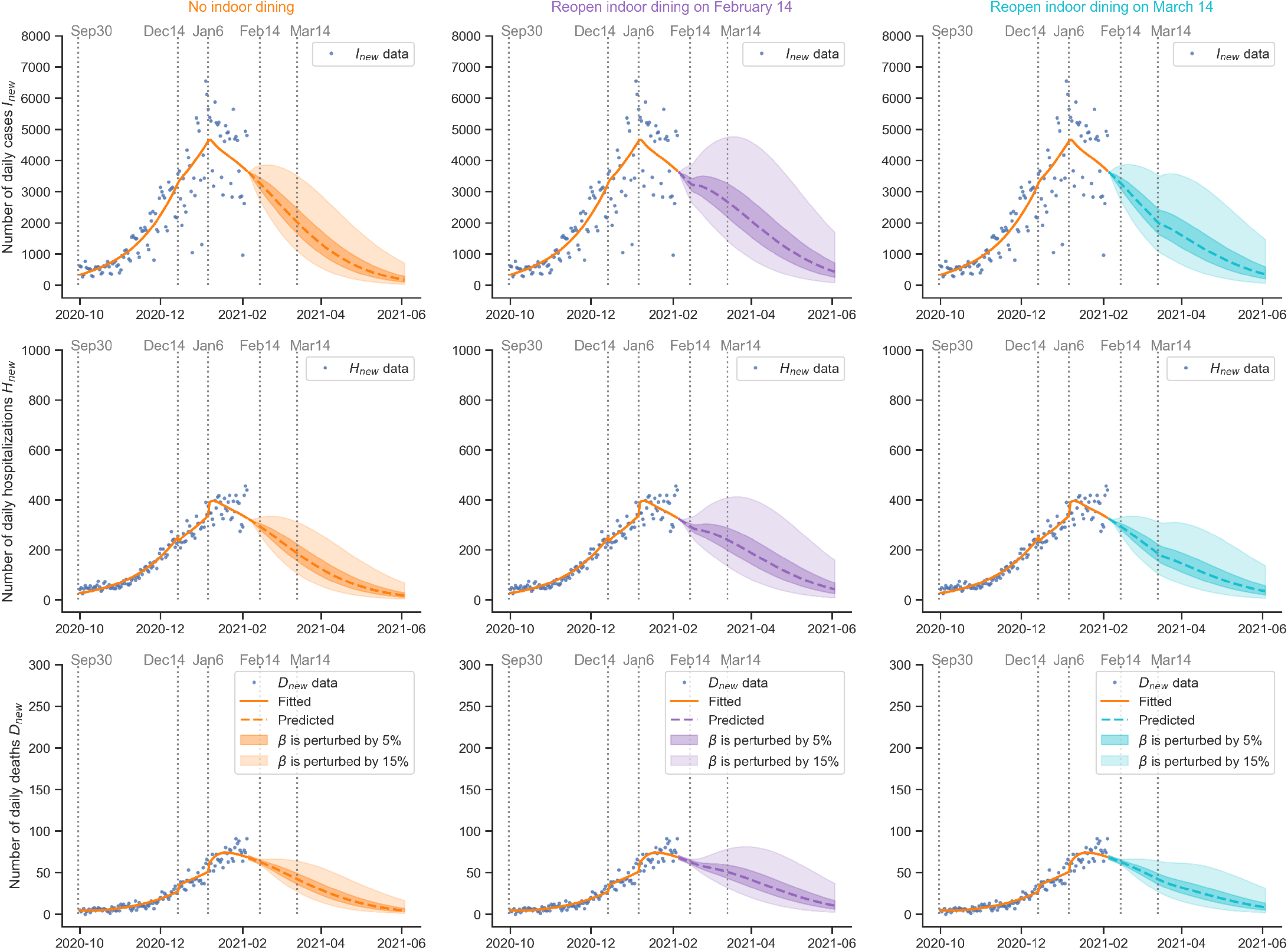
Forecast of daily cases, hospitalizations, and deaths in New York City with uncertainties and scenarios. Reopening scenarios on February 14 and March 14 are considered. An increase in infectious, hospitalized, and deceased population is expected if the restaurants are reopened in the same way as Stage 5 (September 30, 2020 to December 14, 2020). Postponing the reopening of restaurants from February 14 to March 14 may reduce the number of infectious, hospitalized, and deceased individuals. The actual situation might vary depending on the details and implementations of the actual indoor dining policies that take place in 2021.

## Summary

The COVID-19 epidemic is an unprecedented worldwide public health challenge, especially in densely populated areas such as New York City (NYC). Epidemiological models can provide the dynamic evolution of a pandemic but they are based on many assumptions and parameters that have to be adjusted over the time when the pandemic lasts. However, the available data might not be sufficient to identify the model parameters and hence infer the unobserved dynamics. This is typical of any past epidemics or pandemics, and hence a systematic integrated framework is required to make existing or modified models useful for designing health policies.

To this end and after studying the current pandemic for almost a year, we have designed a general framework as shown in Fig 1 for building a trustworthy data-driven epidemiological model, which constructs a workflow to integrate data acquisition and event timeline, model development, identifiability analysis, sensitivity analysis, model calibration, model robustness analysis, and forecasting with uncertainties in different scenarios. The proposed general framework can provide guidance on how to build the appropriate epidemiological model based on the available data in a specific region. The proposed framework can help to assess the structural and practical identifiability of model parameters so that model parameters can be uniquely estimated, and the calibrated model can make more robust and reliable predictions that can be used for evaluating the effect of vaccination and various other scenarios.

In particular, we apply this framework to first endow the SEIR model with more compartments, and subsequently we extend to include vaccination, with the objective of forecasting the transmission dynamics of COVID-19 in NYC under vaccination and different safety measures relaxation scenarios. Based on the proposed general framework, we first acquire data from the NYC’s government’s website and look for major intervention events that could affect the transmission dynamics of the pandemic. We then develop a mathematical model that describes the COVID-19 infection’s biological characteristics by extending the SEIR model to include presymptomatic, asymptomatic, isolated, hospitalized, and deceased individuals. This model takes advantage of all the epidemiological data available from the COVID-19 outbreak in NYC by fitting hospitalizations and disease-related deaths in addition to the daily cases. Furthermore, we incorporate the effects of intervention strategies in the outbreak’s evolution by including time-dependent parameters to capture these variations.

Given a model and epidemiological data, this framework addresses the problem of identifying which parameters can be inferred accurately. We perform two types of identifiability analysis, structural identifiability and practical identifiability analysis, to address this problem. From the structural identifiability analysis, we conclude that five parameters (*β, p, q, ϵ, δ*) of the proposed model are structurally globally identifiable when daily cases (*I*_*new*_), hospitalizations (*H*_*new*_) and deaths (*D*_*new*_) are provided, which is the case for the NYC dataset. However, when one or two observables are not available, one cannot get a trustworthy estimation of these parameters since at least one of the model parameters would be non-identifiable. For the purpose of reliable parameter estimation, one should utilize all the provided data in NYC.

The Fisher correlation matrix method enables us to determine that two out of five parameters need to be fixed due to practical non-identifiability, even if all the data (*I*_*new*_, *H*_*new*_, *D*_*new*_) in the NYC dataset are given. Therefore, we use the values of *δ* and *E* provided by the CDC pandemic planning scenarios, and once we fix these two parameters (*ϵ, δ*), the other three parameters are practically identifiable. For some other cities, however, it can be challenging to maintain careful records of infected, hospitalized, and deceased individuals in an ongoing epidemic. The robustness analysis allows us to conclude that we can still predict some variables with a degree of accuracy despite missing data since in some cases the non-identifiable parameters are not sensitive to the model outputs.

As a result, we fit three parameters (*β, p, q*) given three observables (*I*_*new*_, *H*_*new*_, *D*_*new*_). Sensitivity analysis demonstrates that the parameter *β* is the most important parameter for the prediction of all *I*_*sum*_, *H*_*sum*_, and *D*_*sum*_. Since *p* and *q* do not contribute to *I*_*sum*_, our model may predict *I*_*sum*_ even if *p* and *q* were inaccurate.

Similarly, our model may predict *H*_*sum*_ even if *q* were inaccurate.

We observe that the proposed data-driven epidemiological model can uniquely estimate the model parameters, and the predicted daily cases, hospitalizations, and deaths match well with the available data from NYC’s government’s website. In addition, we employ Monte Carlo simulations to quantify the uncertainties in the parameters and forecast under uncertainties. We employ the calibrated data-driven model to study the effects of the timing of reopening indoor dining. The prediction results indicate that postponing the reopening of restaurants from February 14 to March 14 may reduce the number of infectious, hospitalized, and deceased individuals. Such forecasts can be readily be updated as new data are accumulated. The actual situation might vary depending on the details and implementations of the actual indoor dining policies that take place in 2021 and corresponding updates are required.

## Data Availability

The datasets generated during and/or analysed during the current study are available from the corresponding author on reasonable request.

## Acknowledgments

We gratefully acknowledge the support from ARO/MURI grant W911NF-15-1-0562.

## Author contributions

**Conceptualization:** Sheng Zhang, Joan Ponce, Guang Lin, George Karniadakis

**Data curation:** Sheng Zhang

**Formal analysis:** Sheng Zhang, Zhen Zhang

**Funding acquisition:** Guang Lin, George Karniadakis

**Investigation:** Sheng Zhang, Joan Ponce, Zhen Zhang

**Methodology:** Sheng Zhang, Joan Ponce, Zhen Zhang

**Project administration:** Sheng Zhang, Joan Ponce, Guang Lin, George Karniadakis

**Resources:** N/A

**Software:** Sheng Zhang, Zhen Zhang

**Supervision:** Guang Lin, George Karniadakis

**Validation:** Sheng Zhang, Zhen Zhang

**Visualization:** Sheng Zhang, Zhen Zhang

**Writing — original draft preparation:** Sheng Zhang, Joan Ponce, Zhen Zhang

**Writing — review & editing:** Sheng Zhang, Joan Ponce, Zhen Zhang, Guang Lin, George Karniadakis

## Supporting information

### S1 Text. Model development

We adopt the following step-by-step procedure to build our model:

1. Choose components of the dynamics to simulate the transmission of the disease: susceptible (*S*), exposed (*E*), presymptomatic (*P*), symptomatic (*I*), asymptomatic (*A*), hospitalized (*H*), isolated (*Q*), deceased (*D*), and recovered (*R*).
2. Identify the flow between the components and link them with directed arrows. Assign the rate of the flow with one parameter per arrow except for *S →E*, which is defined by the transmission of the disease. See S1 Fig.
3. Change parameters such that every parameter has physical meaning. We use nine parameters (*p, q, δ, d*_*E*_, *d*_*P*_, *d*_*I*_, *d*_*A*_, *d*_*H*_, *d*_*Q*_) to replace (*w*_1_,, *…w*_9_). See Eq. (13) for the formulas. Note that there is a one-to-one correspondence between the nine parameters we use and (*w*_1_, …, *w*_9_).
4. Add vaccination dynamics *S → R*.

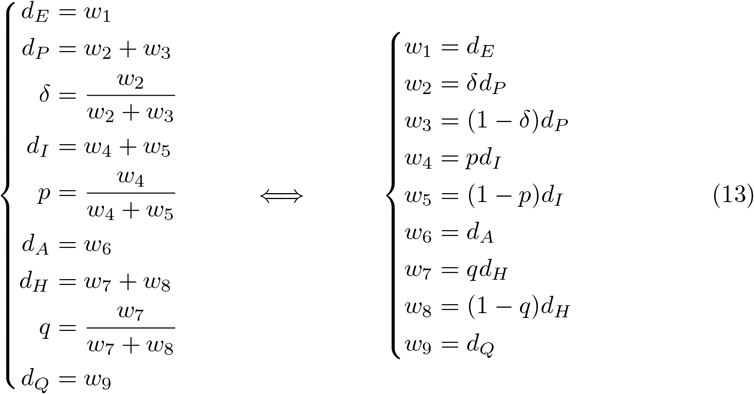

### S2 Text. Parameter settings

Parameter values for the fixed parameters are summarized in Table 2. The percentage of infected people who never show the disease’s symptoms is extracted from the CDC’s current best estimate for this value [38]. The parameter *δ* is obtained by averaging the lower and upper bounds of the estimates for this value from different sources:

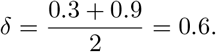

We use data from the hospitalization surveillance network used by the CDC to estimate the median number of days an individual spends hospitalized due to the disease [58]. The numbers provided are specific to individuals admitted to the ICU and not admitted to the ICU divided into age groups; therefore, we perform a weighted average of those values considering the demographic composition of NYC. According to S1 Table, the average number of days of hospitalization for individuals aged 18-49 years is:

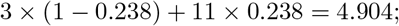

average number of days of hospitalization for individuals aged 50-64 years:

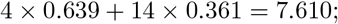

average number of days of hospitalization for individuals aged *≥* 65 years:

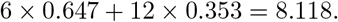

As a result, the total average number of days of hospitalization *d*_*H*_ for all age groups can be obtained in the following way:

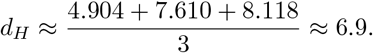

### S3 Text. Basic and control reproduction number of the model in Fig3

The reproduction number in Eq. (5) is obtained using the next generation matrix operator described in [59]. The vector of infected state variables without considering hospitalization is denoted by **x** = (*E, P, I, A*). We define 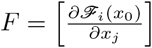 and 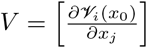, where 𝒻_*i*_ is the rate of appearance of new infections in the *i*th compartment; 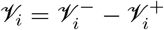, where 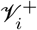 is the rate of transfer of individuals into the *i*th compartment by all other means except for infection, and 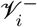 is the rate of transfer of individuals out of the *i*th compartment; and

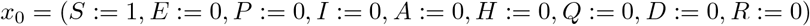

is the disease–free equilibrium state of the system. We have

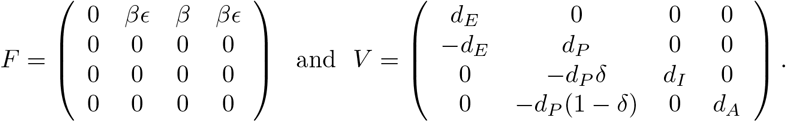

Then,

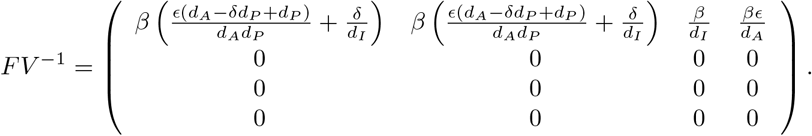

Let *ρ*(*FV* ^*−*1^) denote the dominant eigenvalue of *FV* ^*−*1^. The control reproduction number is given by *ℛ*_*c*_ = *ρ*(*FV* ^*−*1^).

### S4 Text. Definition and algorithm for identifiability analysis

#### Structural identifiability

Suppose we are give a dynamical system of the following abstract form

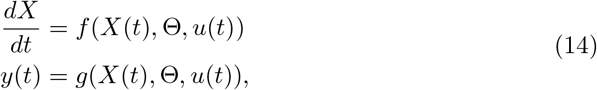

where *X* = (*X*_1_, …, *X*_*n*_) represents the state variables, *y* = (*y*_1_, …, *y*_*m*_) represents the observables, Θ = (*θ*_1_, …, *θ*_*k*_) contains the parameters to identify, and *u*(*t*) represents the input variable to the system. A parameter set Θ is called structurally identifiable if

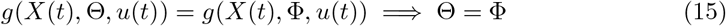

for every Φ = (*φ*_1_, *…, φ*_*k*_) in the same space as Θ. A single parameter *θ*_*i*_ ∈Θ is called structurally identifiable if

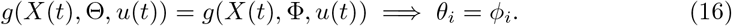

The structural identifiability, defined above, is referred to as global identifiability in [51–54]. Global identifiability is stronger than the so-called local identifiability, which only requires Eq. (15) or Eq. (16) to hold in a neighbourhood 𝒩 (Θ) of Θ. Structural identifiability analysis is usually conducted before the fitting of the model and it does not rely on any data.

We perform the structural identifiability analysis using the software SIAN [56], which is based on differential algebra and Taylor series expansion. Detailed documentations on the theory behind the software can be found in [60], and information regarding the algorithm can be obtained from [56].

SIAN is a randomized algorithm that requires users to choose a hyperparameter that defines the probability of correctness. In this paper we choose 0.999. The algorithm first replaces the observables with their truncated Taylor series and reduces the identifiability problem to the problem of studying the generic fiber of the map between the parameter space and the space of truncated observables, which is a map between finite-dimensional varieties. Due to the hardness of analyzing the generic fiber, a step to simplify computation is applied in the algorithm by studying fiber at a specific point instead of generic ones. The correctness of this step is controlled by the hyperparameter aforementioned. The problem is then turned into checking the consistency of a system of algebraic equations. The Buchberger algorithm allows the computation of the Grobner basis of the system. This basis shows whether the algebraic relations’ parameters have a single solution/finitely many solutions/infinitely many solutions, which implies the model would be structurally globally identifiable/locally identifiable/not identifiable.

### Practical identifiability and correlation matrix

A parameter is called practically identifiable if it can be uniquely determined from the discrete data points instead of the structure of the equation. A structurally identifiable parameter may not be practically identifiable given the noise and limited measure points in the data. Unlike structural identifiability, there is no consistent definition of practical identifiability in the literature. Nevertheless, several methods including correlation matrix methods have been proposed to describe practical identifiability quantitatively [19, 57].

Statistical correlation can be used to describe some of the practical non-identifiability phenomena. Specifically, one can use the Fisher Information Matrix (FIM) to calculate the correlation matrix of Θ in Eq. (14). FIM is defined as:

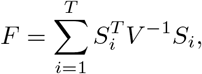

where *V* is covariance matrix of the measurements error. 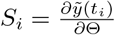 is the sensitivity matrix at time *t*_*i*_, which can be calculated by solving the adjoint equation:

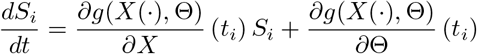

using automatic differentiation libraries. It is proved that the correlation matrix is equal to *F* ^*−*1^ by the Cramér–Rao theorem [61]. Once the correlation matrix is obtained, one can determine whether a parameter is identifiable by checking whether it is correlated with the other parameters. If *θ*_*i*_ is strongly correlated with *θ*_*j*_, then neither of these parameters is practically identifiable because they cannot be uniquely determined from the data.

### S5 Text. Definition and algorithm for model robustness

The procedure of Monte Carlo simulation for model robustness can be summarized into the following steps:

- Solve Eq. (14) with Θ equal to the estimated value 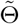 to get the calibrated observables 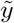 at the time stamps where the original data are sampled. For example, for NYC dataset in the fully-observed case, we assume 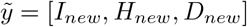, i.e., the daily increase in *I*_*sum*_, *H*_*sum*_, and *D*_*sum*_.
- Multiply the calibrated observables 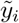 by independent and identically distributed Gaussian random noise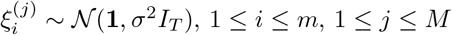, where *T* is the number of time stamps, *I*_*T*_ is the *T ×T* identity matrix, and *M* is the number of Monte Carlo steps. By this method we generate a dataset of size *M* :

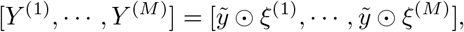

where is the elementwise product and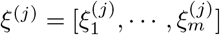.
- The parameters are estimated again using these perturbed observables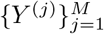. The estimated value for the *i*th unknown parameter *θ*_*i*_ using *Y* ^(*j*)^ is denoted as 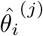. The sensitivity-based average relative error (SARE) for *θ* is defined as:

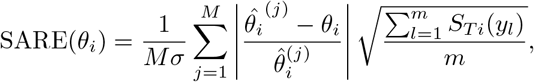

where *S*_*T i*_(*y*_*l*_) is the total-order sensitivity, which is the contribution to the variance of *y*_*l*_ by *θ*_*i*_. In other words, it quantifies the influence of *θ*_*i*_ on the output variable. For more important parameters which have a higher *S*_*T i*_, we require more robustness. This is taken into account by the multiplier 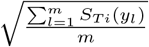 in the definition of SARE. Finally, we define the maximum sensitivity-based average relative error (MSARE) of the model to be the largest SARE across all the model parameters:

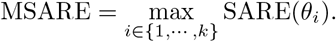

If MSARE *<* 1, we say that the model is robust to perturbation.

**S1 Table.**
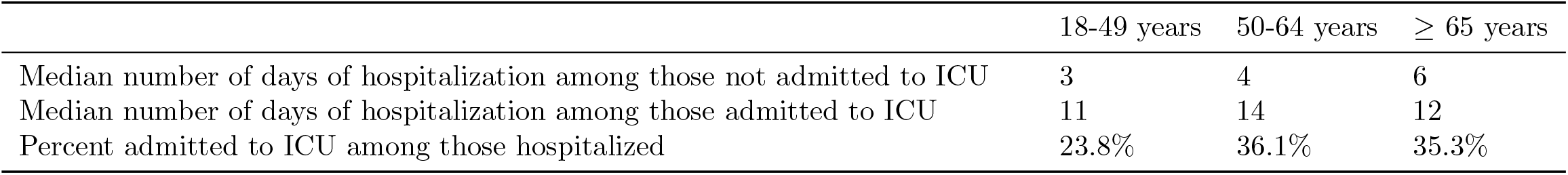
Data from CDC website (COVID-19 Pandemic Planning Scenarios) to estimate *d*_*H*_.

| | 18-49 years | 50-64 years | $\geq 65$ years |
| --- | --- | --- | --- |
| Median number of days of hospitalization among those not admitted to ICU | 3 | 4 | 6 |
| Median number of days of hospitalization among those admitted to ICU | 11 | 14 | 12 |
| Percent admitted to ICU among those hospitalized | 23.8% | 36.1% | 35.3% |

**S2 Table.** Structural identifiability of the model with *H* as an observable. Structural identifiability is tested when *H* instead of *H*_*sum*_ is given as an observable. It turns out that the identifiability result is similar to Table 3.

| Parameter | $I_{sum}, H, D_{sum}$ | $I_{sum}, H$ | $H, D_{sum}$ | $H$ |
| --- | --- | --- | --- | --- |
| $\beta$ | global | global | not | not |
| $p$ | global | global | not | not |
| $q$ | global | not | global | not |
| $\epsilon$ | global | global | global | global |
| $\delta$ | global | global | global | global |

**S1 Fig.**
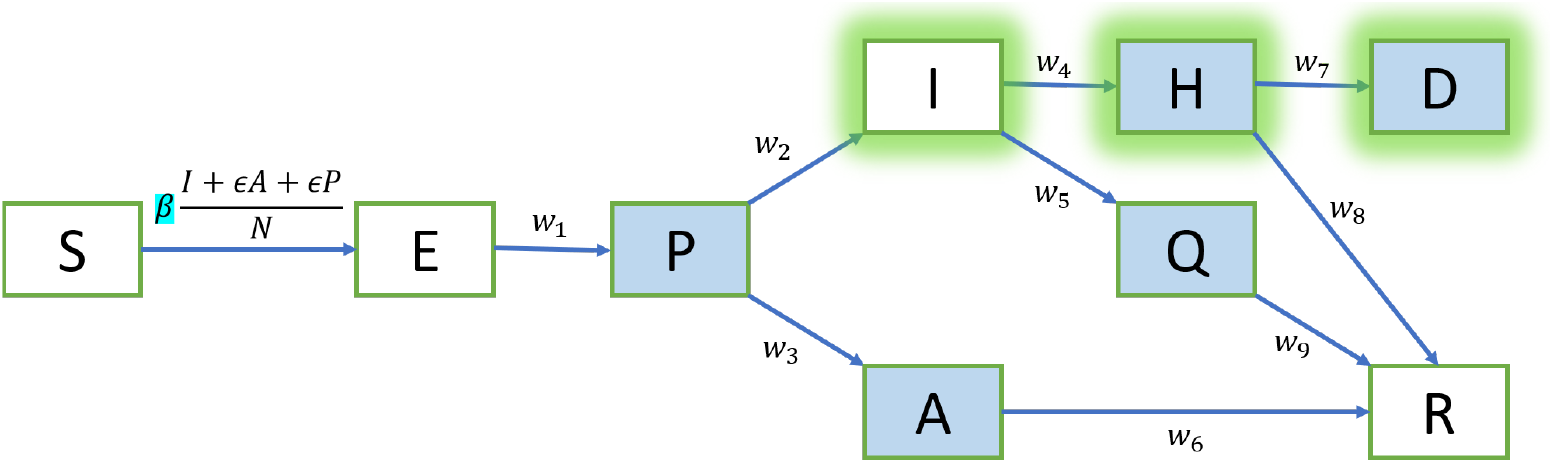
Raw model. The graph demonstrates an intermediate step of building the proposed model in Fig 3. It has the same compartmental structure as the final model but the inflows and outflows of different compartments are parameterized by unknown *w*_*i*_’s. It can be transformed to the final model through a change of variable in Eq. (13) and a supplement of vaccination dynamics.

**S2 Fig.**
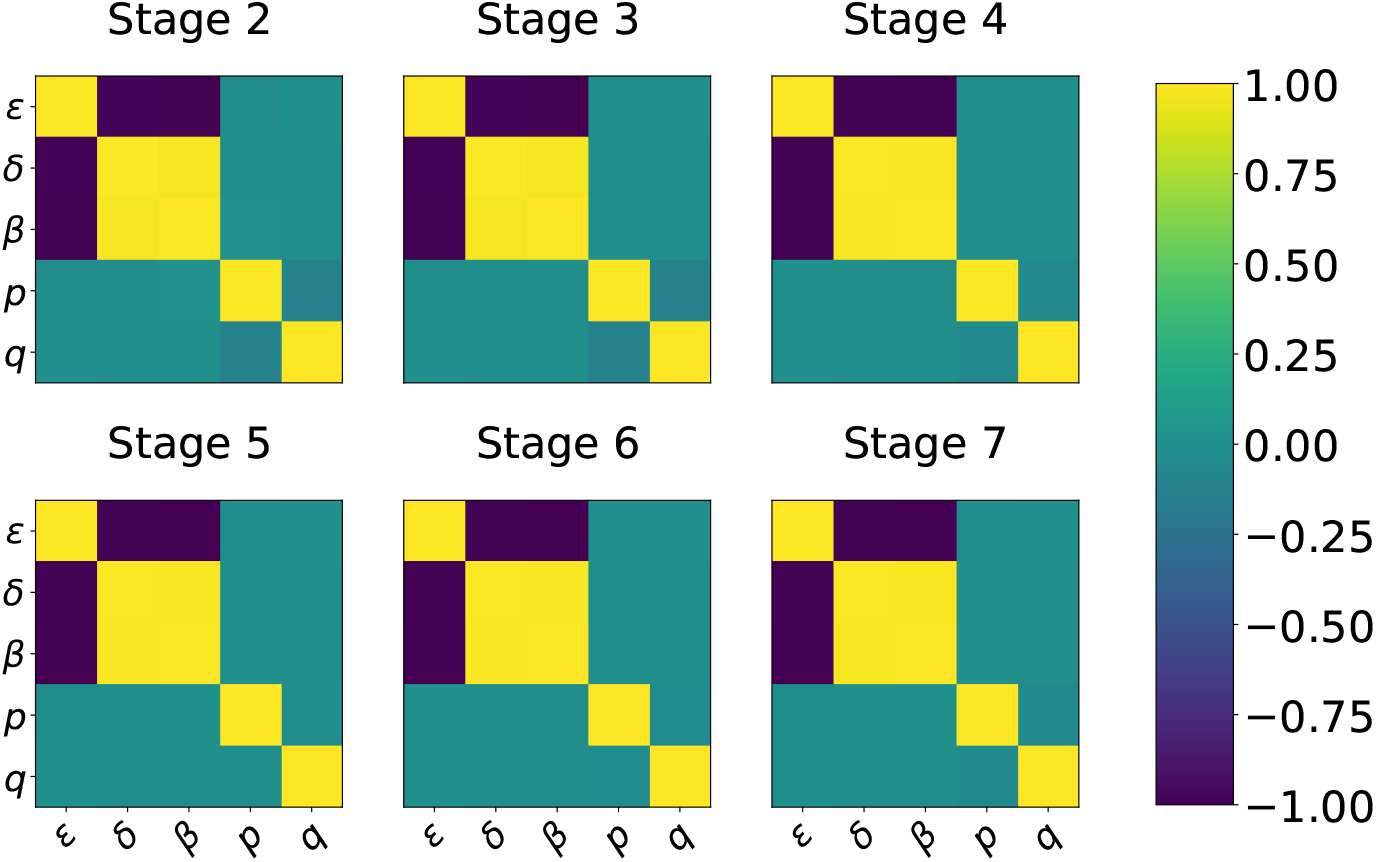
Correlation matrix of *β, p, q, δ*, and *E* in Stage 2 to Stage 7. Green means (almost) not statistically correlated while yellow/purple represents positively/negatively correlated. The correlation matrices are similar in theses stages, i.e., there is always correlation between *β, E, δ* while there is no correlation between *p, q* and the other parameters.

**S3 Fig.**
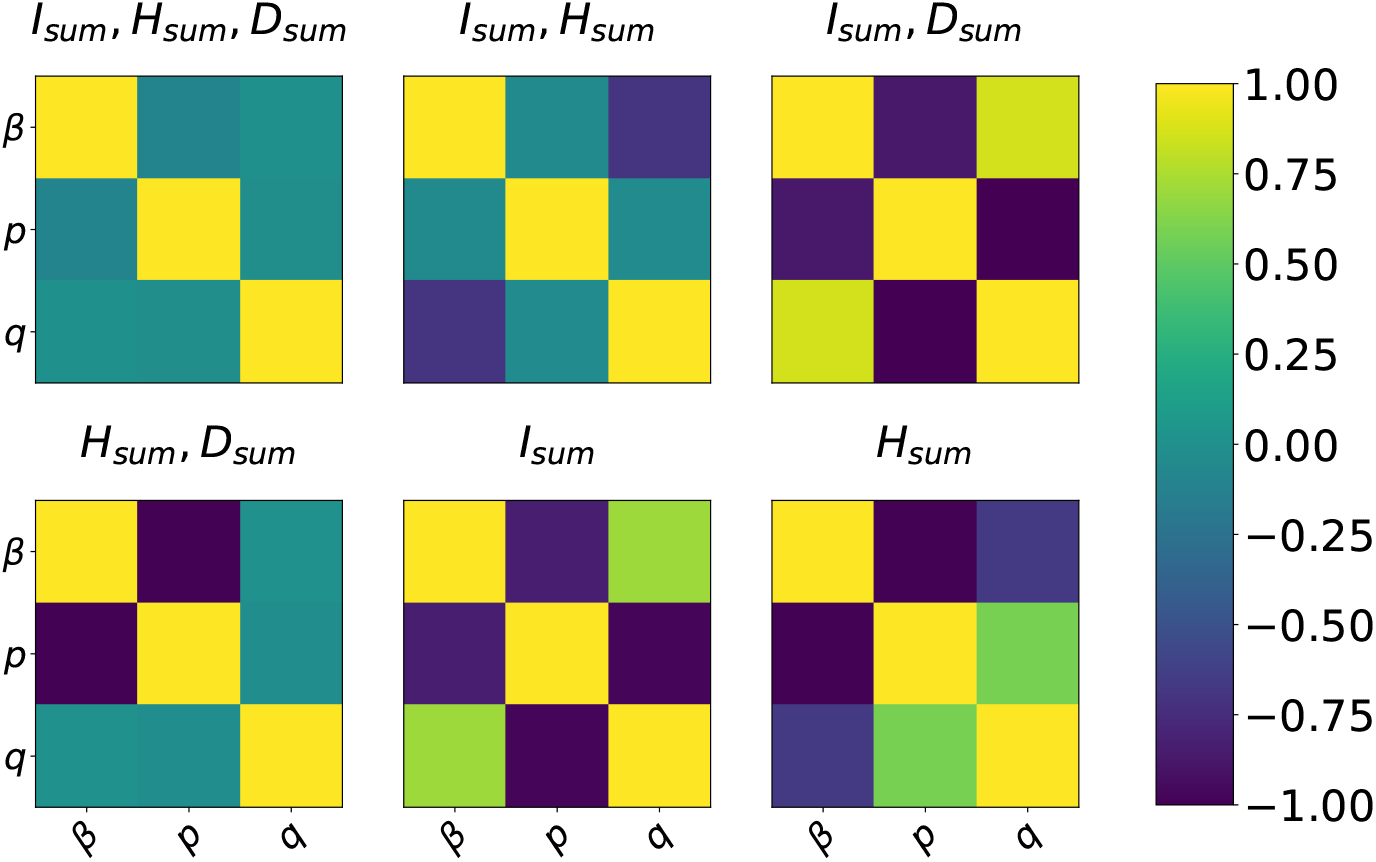
The correlation matrix of *β, p*, **and** *q* **in the setting of different observables, calculated in Stage 1**. Green means (almost) not statistically correlated while yellow/purple represents positively/negatively correlated. When *I*_*sum*_, *H*_*sum*_, and *D*_*sum*_ are available, the model is practically identifiable. In other cases, there is correlation between the parameters.

**S4 Fig.**
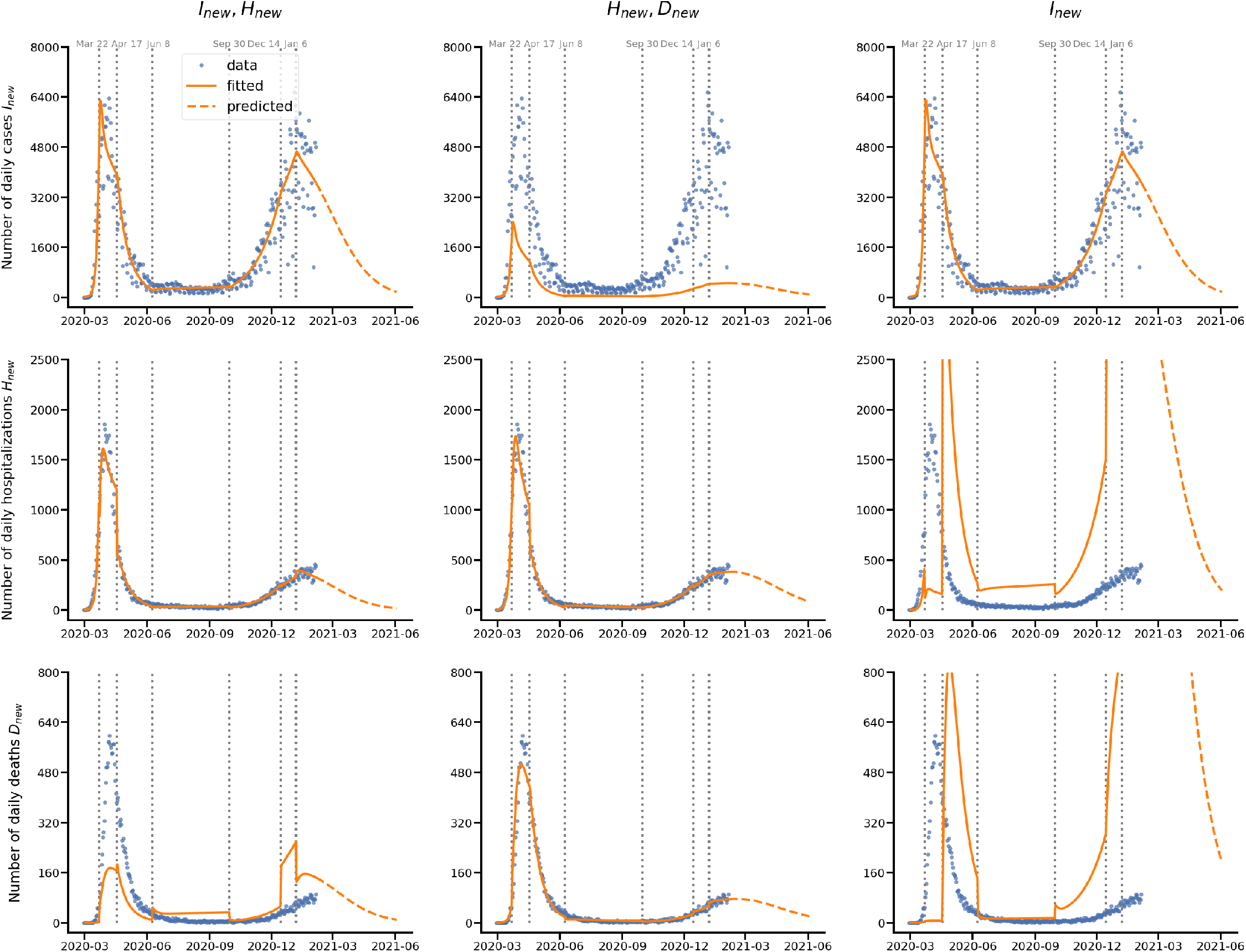
Fitting and prediction with the data given as. (*I*_*new*_, *H*_*new*_), (*H*_*new*_, *D*_*new*_), **or** (*I*_*new*_). Other settings are the same as Fig 6. Each row represents the fitting and prediction of an observable. Each column represents an observable setting. When only *I*_*new*_ and *H*_*new*_ are available, the fitting and prediction for *D*_*new*_ is inaccurate, while the fitting and prediction for *I*_*new*_ and *H*_*new*_ are correct. When only *H*_*new*_ and *D*_*new*_ are available, the fitting and prediction for *I*_*new*_ is inaccurate, while the fitting and prediction for *H*_*new*_ and *D*_*new*_ are correct. When only *I*_*new*_ is given, the model can only predict *I*_*new*_ correctly. These results can be explained by model robustness analysis.

**S5 Fig.**
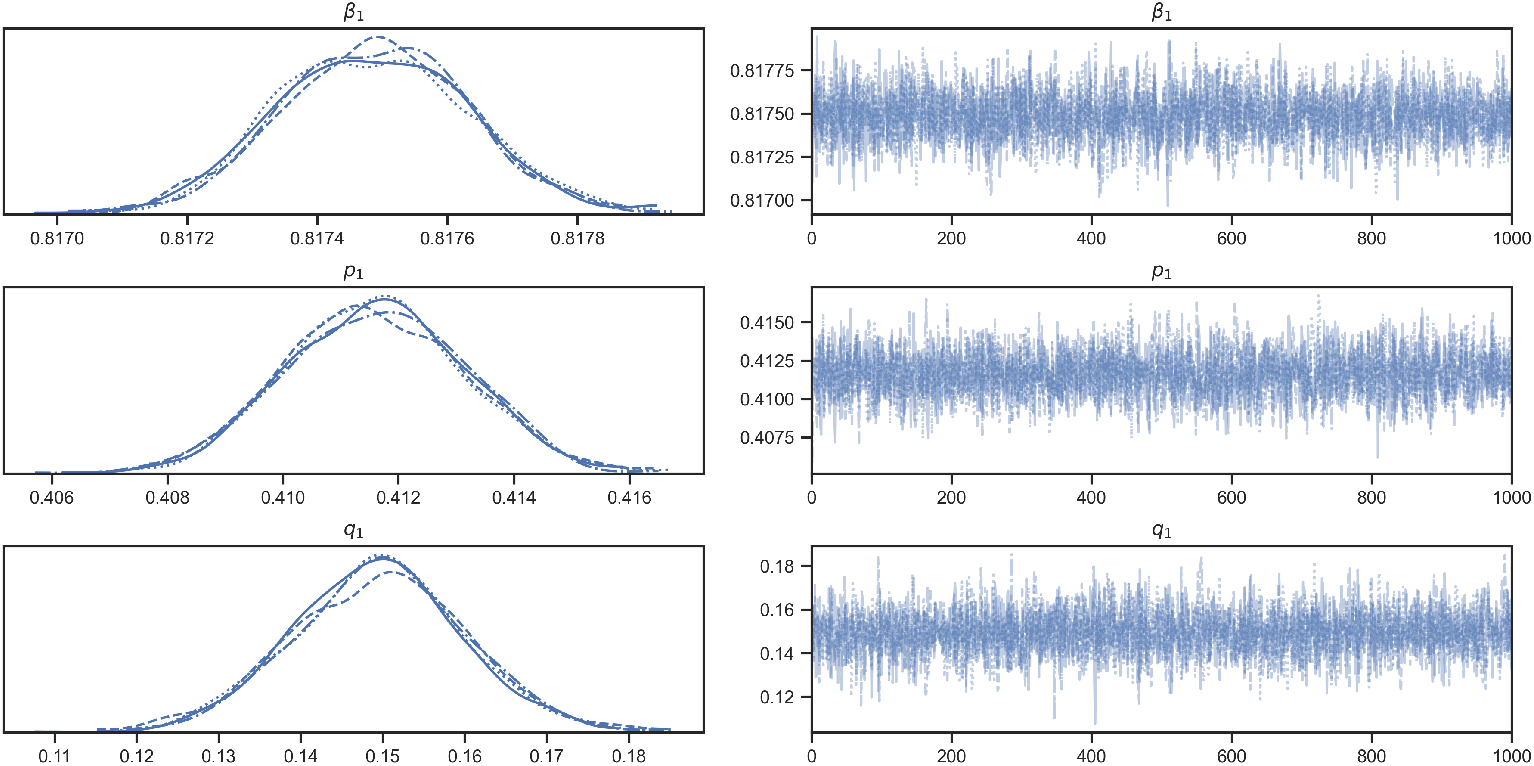
MCMC simulation in Stage 1. Four chains of 1000 samples are drawn with 200 burn-in samples in every chain. **[Left Column]** Posterior distributions of the parameters (*β, p, q*). The narrow posterior distributions indicate that our numerical algorithm is robust, the quantity of data is sufficient for our approach of minimizing the loss function Eq. (10), and the parameters are indeed close to constant. **[Right Column]** Sampling processes of the parameters. We can see that the chains are well-mixed, which implies the convergence of the sampling.

**S6 Fig.**
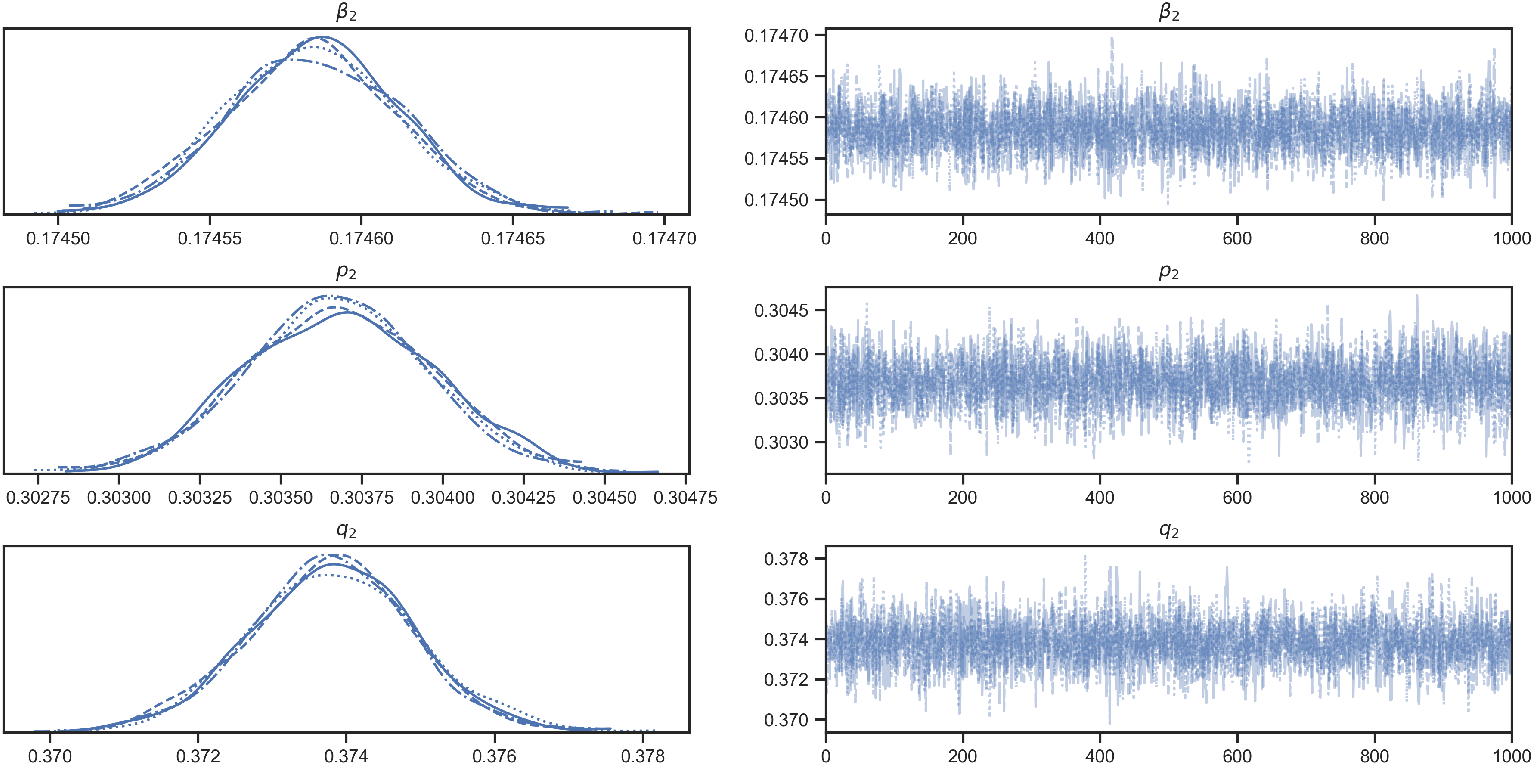
MCMC simulation in Stage 2. Other settings are the same as S5 Fig.

**S7 Fig.**
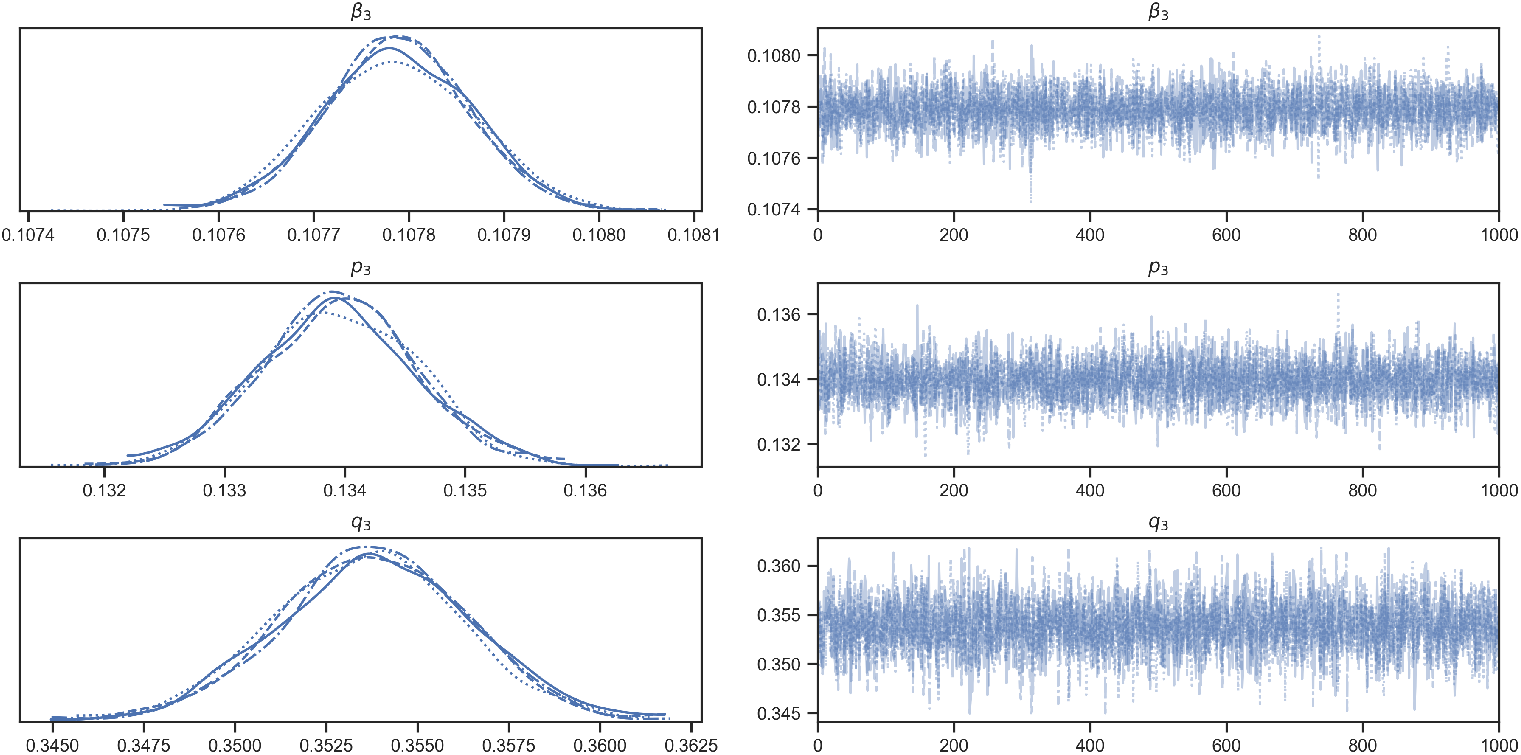
MCMC simulation in Stage 3. Other settings are the same as S5 Fig.

**S8 Fig.**
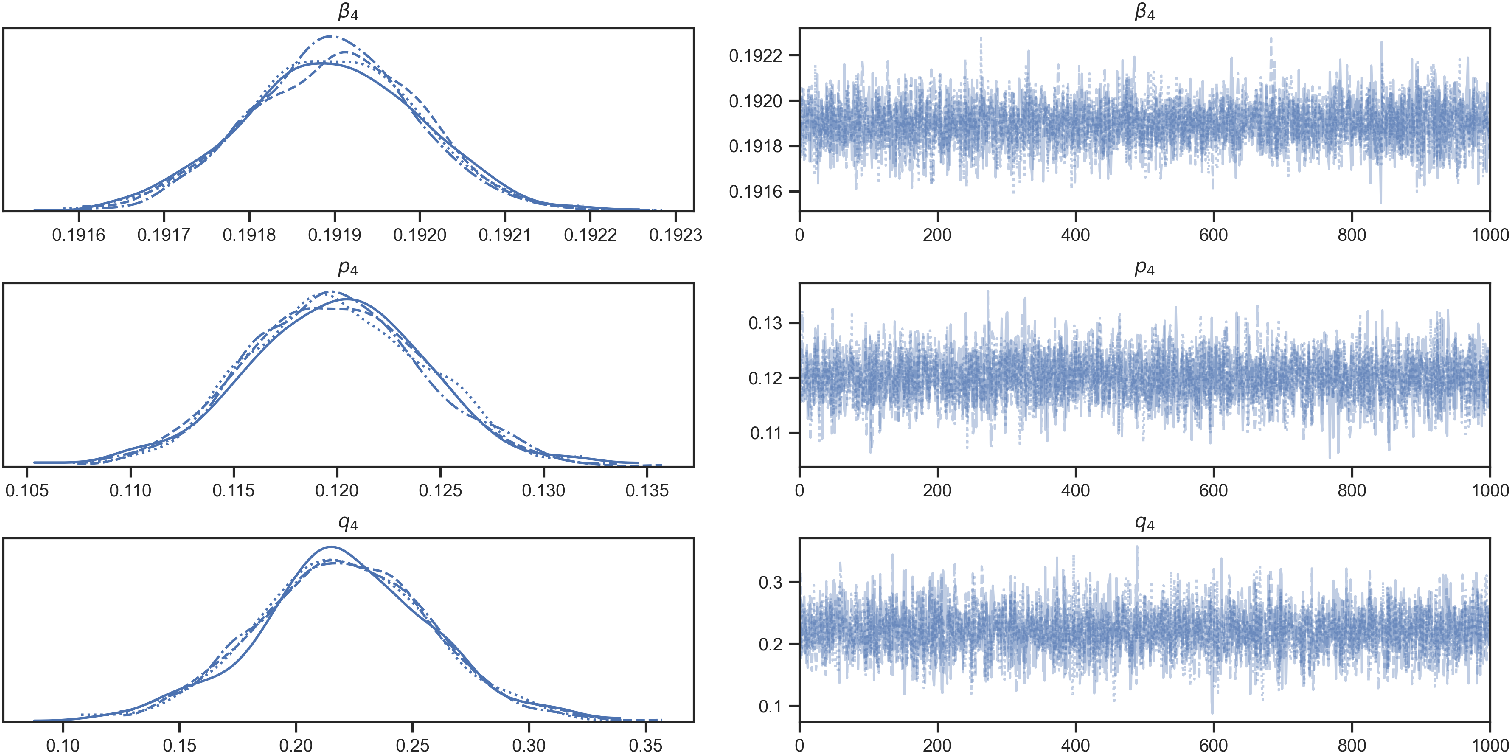
MCMC simulation in Stage 4. Other settings are the same as S5 Fig.

**S9 Fig.**
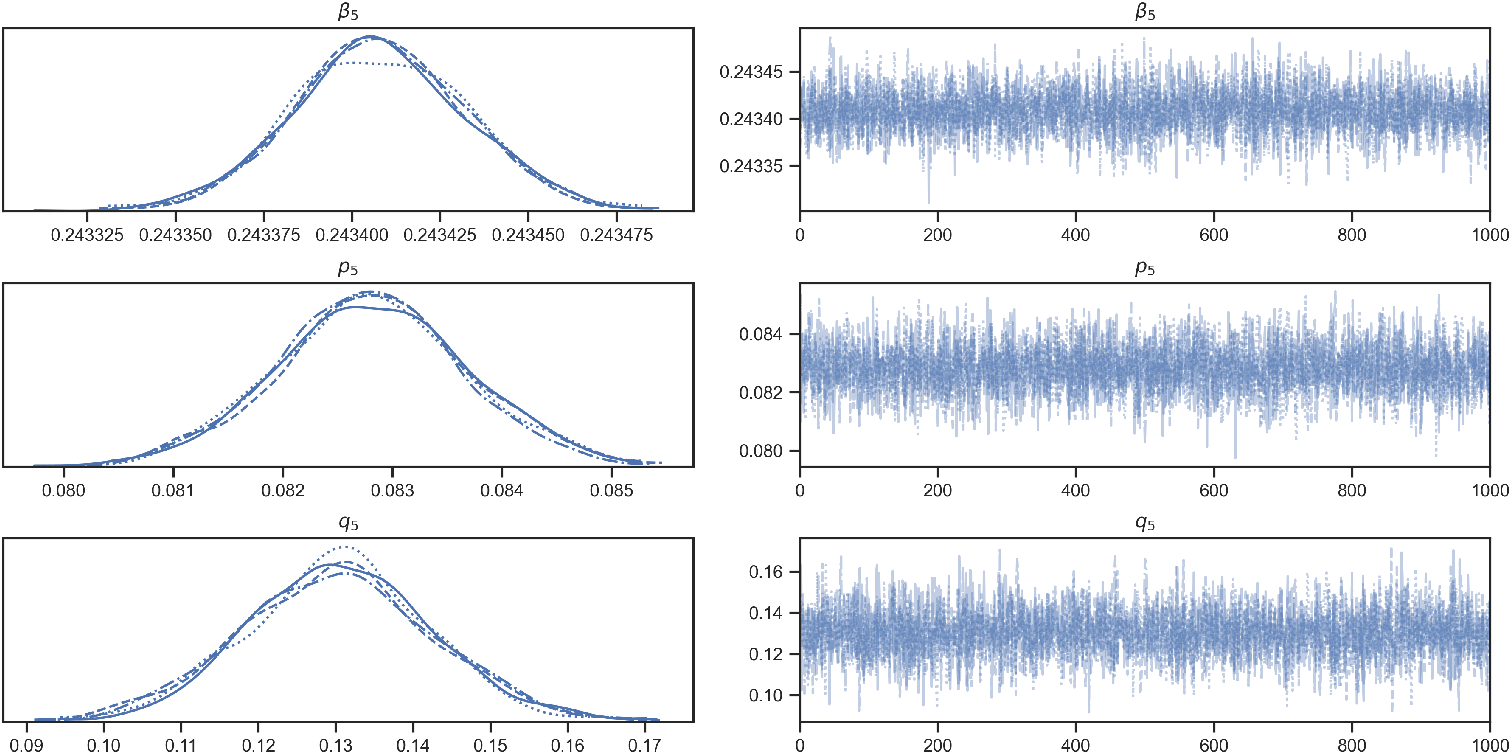
MCMC simulation in Stage 5. Other settings are the same as S5 Fig.

**S10 Fig.**
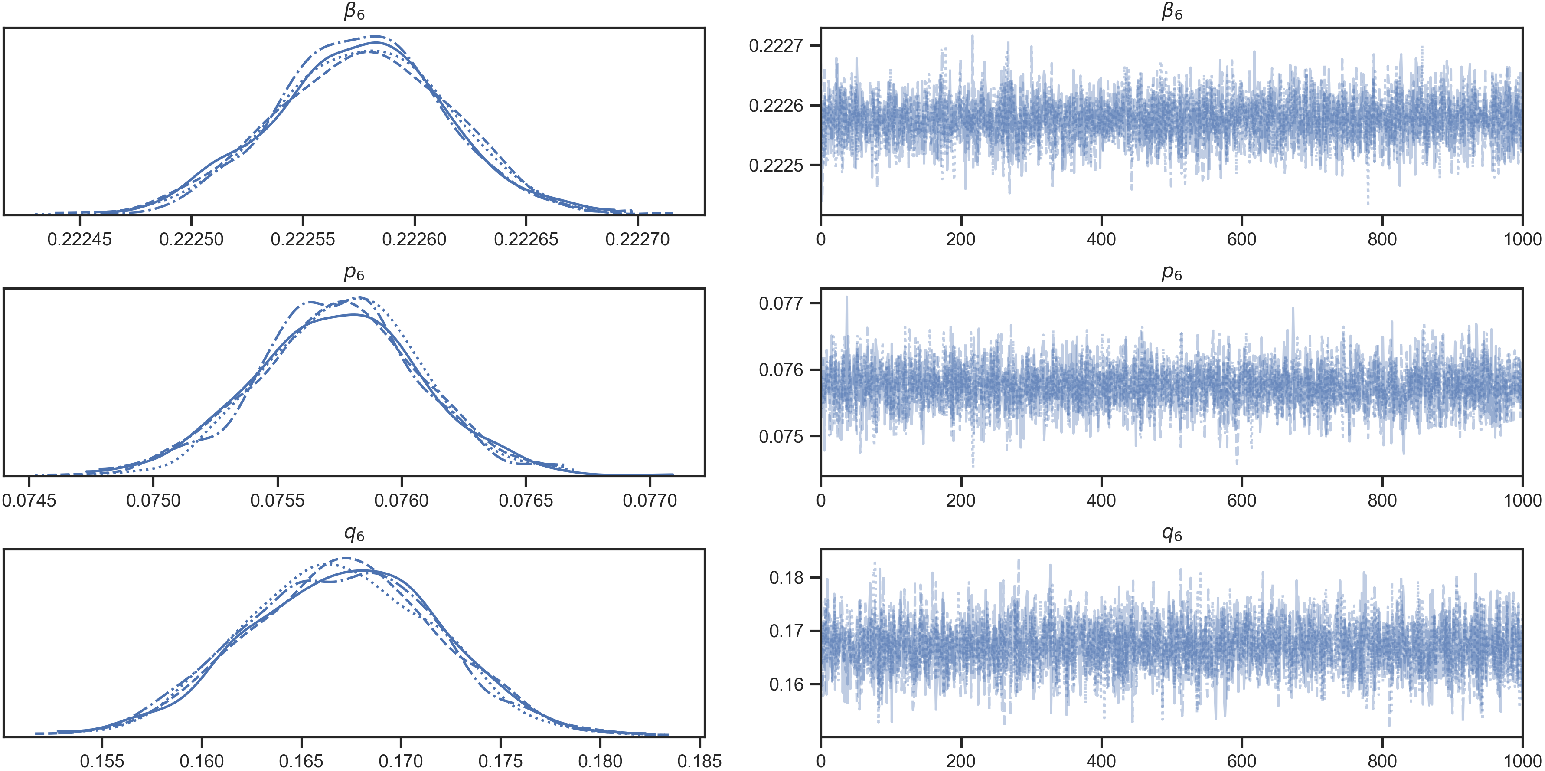
MCMC simulation in Stage 6. Other settings are the same as S5 Fig.

**S11 Fig.**
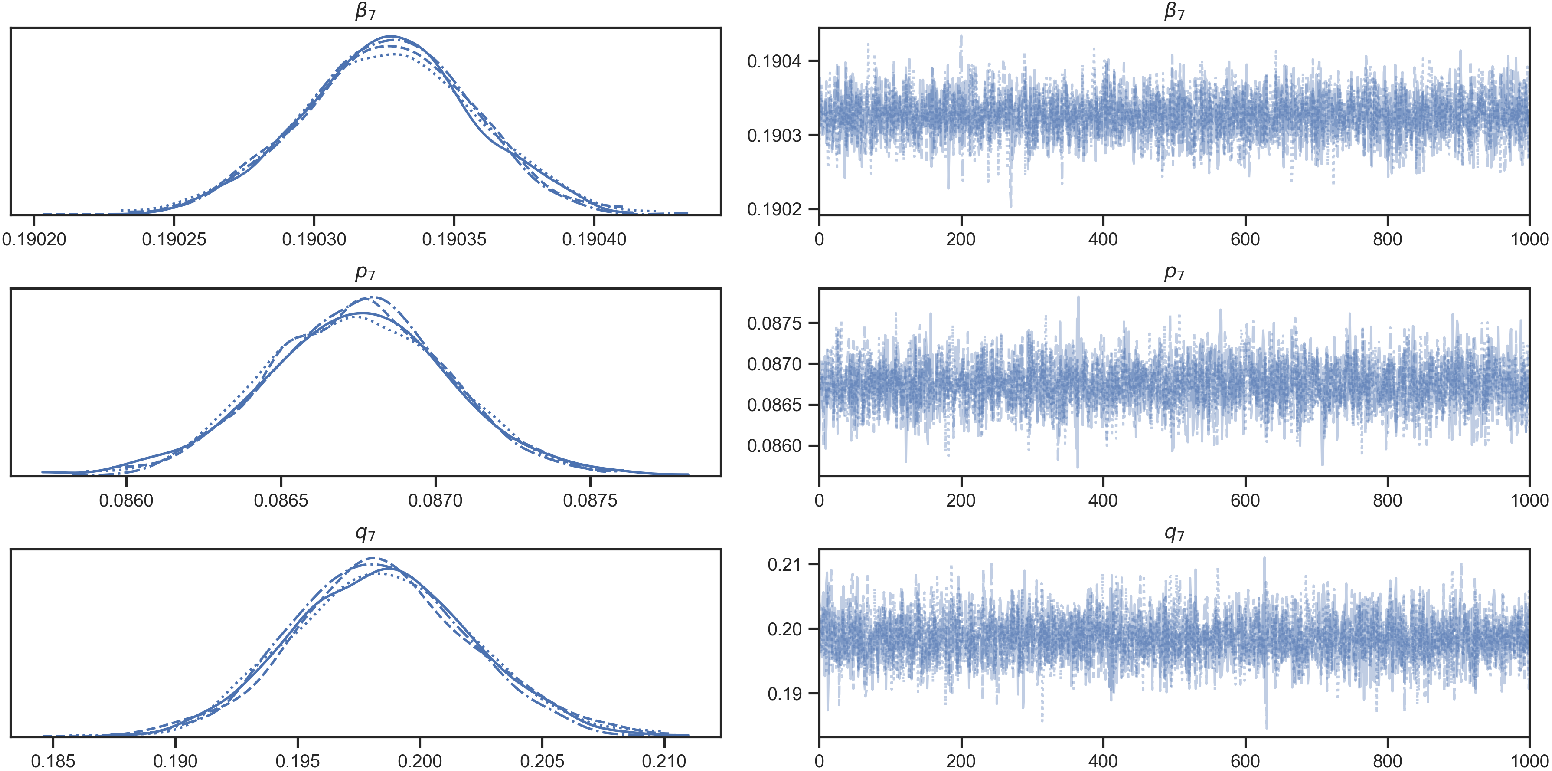
MCMC simulation in Stage 7. Other settings are the same as S5 Fig.

